# A Network Meta-Analysis of Quality of Life in Heart Failure with Reduced Ejection Fraction

**DOI:** 10.1101/2025.03.12.25323680

**Authors:** Robert Margaryan, Nariman Sepehrvand, Wouter Ouwerkerk, Jasper Tromp, Ricky D. Turgeon, Justin A. Ezekowitz

## Abstract

**Background:** While the effect of various combinations of treatments on mortality and morbidity outcomes in heart failure with reduced ejection fraction (HFrEF) have been evaluated, the impact on quality of life is unknown. This study evaluated and compared the composite impact of pharmacological therapies on quality of life in HFrEF using a frequentist network meta analysis and systematic review methodology.

**Methods:** We searched MEDLINE, EMBASE and Cochrane Central Register of Controlled Trials for randomized controlled trials published between January 1987 - August 2024. We included all contemporary and efficacious HFrEF therapies used in adults. The primary outcome was the mean change in QoL score measured through the Kansas City Cardiomyopathy Questionnaire and the Minnesota Living with Heart Failure Questionnaire, expressed as mean difference (MD).

**Results:** We identified 41 randomized controlled trials representing 41145 patients which had a median of 276 (IQR 105, 464) participants who were mostly male (76.5%) with a mean left ventricular ejection fraction of 28%, and a median follow up time of 5 months (IQR 3,8). The combinations which resulted in the greatest improvement of quality of life were ARNi + BB + MRA + SGLT2i [MD 7.11 (95% CI -0.99-15.22)], which did not have a statistically significant effect, followed by ARNi + BB + SGLT2i [MD 5.33 (95% CI 0.40-10.25)], ACEi + BB + MRA+ SGLT2i [MD 5.32 (95% CI -2.63-13.26)], ACEi + BB + MRA + ivabradine [MD 5.24 (95% CI -3.07-13.55)]. Individually, the treatments which led to the greatest improvement in quality of life were H-ISDN [MD 3.87 (95% CI, -0.73 to 8.47)], which did not have a statistically significant effect, followed by SGLT2i [MD 3.37 (95% CI 1.44-5.30)], ivabradine [MD 3.26 (95% CI 0.08-6.43)], ARNi [MD 2.62 (95% CI -3.24-8.47)] and vericiguat [MD 1.00 (95% CI -3.18-5.18)].

**Conclusion:** A composite of ARNi + BB + MRA + SGLT2i or ARNi + BB + SGLT2i was the most effective at improving quality of life in patients with HFrEF.

## Introduction

Heart failure (HF) affects more than 50 million people globally and is associated with frequent hospital and emergency department visits, reduced health-related quality of life (HRQoL) and high mortality rates.^1–5^ There are a growing number of pharmacological treatments available for heart failure with reduced ejection fraction (HFrEF), such as angiotensin-converting enzyme inhibitors (ACEi), angiotensin receptor blockers (ARB), beta-blockers (BB), mineralocorticoid receptor antagonists (MRAs), digoxin, isosorbide dinitrate-hydralazine (H-ISDN), ivabradine, angiotensin receptor–neprilysin inhibitors (ARNi), sodium-glucose cotransporter-2 inhibitors (SGLT2i), vericiguat, and omecamtiv mecarbil (OM), allowing for differing combinations of medications. Individual medications have been tested additively, with each successive medication demonstrating a reduction in a primary endpoint (typically a decrease in the risk of mortality and hospitalization). Current guidelines recommend the initiation and up titration of an ARNi or ACEi or ARB, together with a BB, MRA and SGLT2i referred to as guideline-directed medical therapy (GDMT).^4,5^

While several systematic reviews and meta-analyses have previously shown the individual effects of various classes of drugs used to treat HFrEF on HRQoL, there has not yet been an assessment which has compared available treatment combinations.^5–20^ As a recent systematic review and network meta analysis of pharmacological treatment effects on patient outcomes in HFrEF has shown evidence for the additive nature of these medications, evaluating the treatment effect of these combinations on HRQoL is feasible.^21^ Since most patients with HFrEF are prescribed medications of various classes, understanding the effect of different combinations and determining the most effective treatment for improving HRQoL would support patients and physicians in achieving patient goals.^22^ Accordingly, we conducted a systematic review and network meta-analysis to estimate and compare the impact of pharmacological therapies, individually and in combination, in patients with HFrEF.

## Methods

### Protocol

The detailed prespecified protocol for this study can be found in the **Supplemental Appendix**.

### Study Design

We performed a systematic review and network meta-analysis using a frequentist framework. The study results are reported according to the PRISMA extension statement for systematic reviews incorporating network meta-analyses.

### Search Strategy, Eligibility and Selection Criteria, and Data Collection

We systematically searched MEDLINE, EMBASE, and the Cochrane Central Register of Controlled Trials for randomized controlled trials published between January 1st, 2021 and August 10th, 2024. Subject headings, MeSH terms, and /or keyword searches were based on the search terms used in a previous network meta-analysis (Tromp and Ouwerkerk et al.) adapted to identify quality of life outcomes.^21^ The full search terms are outlined in the **Supplemental Appendix**. Search results were supplemented with the trials included in Tromp and Ouwerkerk et al. (searched from January 1st, 1987 - January 1st, 2021) and trials included in related meta-analyses by other authors.^21^

We evaluated randomized controlled trials (RCTs) that investigated the effects of GDMTs and other drugs evaluated in patients with HFrEF. Pharmacological agents considered included digoxin, isosorbide dinitrate and hydralazine, ACEi, ARB, BB, MRA, ivabradine, ARNi, SGLT2i, vericiguat, and omecamtiv-mecarbil. Target studies were limited to adults (aged ≥18 years) with HFrEF, enrolled in the outpatient setting or after stabilization following hospitalization for HF who were assessed using the Kansas City Cardiomyopathy Questionnaire (KCCQ) or the Minnesota Living With Heart Failure Questionnaire (MLHFQ) as these are reliable assessment tools which are most prevalent in contemporary clinical trials.^23–25^ Studies were excluded when the entire study population was composed of patients with a concomitant diagnosis that likely had a major effect on HRQoL outcomes of interest such as trials only including patients with diabetes. Studies that only investigated therapies for patients in acute HF episodes, studies that only compared agents within the same drug group, and studies that were not available in English were excluded.

Two investigators (R.M. and N.S.) screened the titles and abstracts of retrieved citrations to identify trial eligibility. The full texts of the trials which were deemed eligible were then independently screened by the same 2 investigators. Discrepancies were resolved by discussion and consensus at each step. Data from the final set of eligible trials after full text review were extracted by the same two authors, with additional review for inconsistencies.

### Outcomes

The primary outcome of interest of the study was the change in health related HRQoL, assessed as the KCCQ Overall Summary Score (OSS) and the MLHFQ Overall score as these are the most comprehensive and commonly recorded score domains. The QoL scores from studies using the KCCQ or MLHFQ were standardized on the KCCQ scale based on a relevant clinical improvement conversion regression formula established by Nassif et al. (MLWHF change = KCCQ OSS change * (-0.74902) – 2.92430).^26^

Using this formula, a common overall score was established and mean differences (MD) for the change in HRQoL were calculated. Results are presented as a MD with 95% confidence intervals (CI). In addition, we analyzed the likelihood of drug discontinuation due to any reason to evaluate previously reported associations between the likelihood of discontinuation and HRQoL experience.^27^

### Network Meta-Analysis

We constructed network meta-analysis models using a frequentist framework through which we generated a random and fixed effects model. We were unable to predict whether there would be a large amount of heterogeneity in the model, therefore, we assessed the appropriateness of both models considering estimates of heterogeneity. A fixed effects model is preferred, but when heterogeneity results are high, the results of the random effects model are the primary results - the results of the fixed effects model are presented in the **Supplemental Appendix**. We assessed the change in HRQoL scores for the individual components of the network compared with placebo and employed an additive component network meta-analysis model to evaluate the influence of individual treatment components, as many treatments were combinations of several common components. This model assumes that the effect of treatment combinations is the sum of the effects of its components, which has been accepted for similar network meta-analyses evaluating HFrEF therapies. Therefore, this network meta-analysis compared different combinations of treatments based on the treatment given and the background therapy within a trial. Treatment combinations that are not included in the network cannot be compared and a connected network is important for accurate results. For a given trial, we considered patients as receiving a background therapy if ≥50% of patients in both arms were on that therapy at baseline. Given the mixed use of ARNi, ACEi, and ARBs in recent clinical trials, it was difficult to accurately place these patients on a background therapy of ARNi, ACEi, or ARB individually while it was clear that the use of ACEi/ARB/ARNi was high. To accurately represent the background therapies of the trial, we considered these trials against a background of ARNi if more than 40% of the patients using ACEi/ARNi/ARB in either arm were on ARNi, if less than 40% were taking an ARNi then the background therapy was considered to be an ACEi.^28–37^ We reported our findings as the MD for HRQoL outcomes. Treatments were ranked using the P-Score, the frequentist equivalent of the surface under the cumulative ranking curve (SUCRA), which shows the proportion of treatments that are worse than the treatment in question.^38^

We used a Cochrane risk of bias assessment tool version 2 (RoB2) to evaluate the internal validity and bias of individual trials.^39^ Using the August 2019 version of the RoB2, Each trial was assessed to determine the presence of bias arising from the randomization process, bias due to deviations from intended interventions, bias due to missing outcome data, bias in the measurement of the outcome, and bias in the selection of the reported results. The risk of bias was classified as low risk, having some concerns, or high risk (major concerns) of bias in each domain using a combination of signaling questions and tool algorithms to inform a within-study risk of bias judgment. All studies were included regardless of their risk of bias to determine the confidence in individual comparisons according to the Grading of Recommendations, Assessment, Development, and Evaluations (GRADE) framework using the Confidence in Network Meta-Analysis (CINeMA).^40^ The CINeMA framework considers 6 domains to assess confidence: 1) within-study bias; 2) reporting bias; 3) indirectness; 4) imprecision; 5) heterogeneity; and 6) incoherence. RoB2 scores were used to estimate within-study bias.

Reporting bias was formally tested using the Egger test, which assesses the symmetry of funnel plots.^41^ When this was found to be nonsignificant, we considered the risk of reporting bias as “low.” Indirectness, which captures transitivity in the network, was assessed as “low” based on published guidelines. Imprecision compares the treatment effects included in the 95% CI with the range of equivalence using clinically important effects, which were considered to be a change in score of 5. To assess heterogeneity, we calculated I^2^ values.^42^ I^2^ values are calculated using the chi-square statistic and its degrees of freedom and represent the amount of inconsistency in the network. Incoherence captures transitivity, which stipulates that two treatments can be compared indirectly via an intermediate treatment node. If estimates from direct and indirect evidence disagree, transitivity does not hold and there is incoherence within the network. Incoherence was assessed through a visual evaluation of differences between evidence based on direct and indirect comparisons and a global test based on a random-effects design-by-treatment interaction model where a nonsignificant *P*-value means that there is no incoherence. In sensitivity analyses, to assess the validity of the standardization of KCCQ and MLHFQ overall scores, we separately analyzed trials that used the MLHFQ using the converted scores and the raw scores and compared these results. The fixed and random effects network meta-analysis was performed using the *netmeta* package in R (R Foundation); *P*-values <0.05 were considered statistically significant. Values that were not reported were imputed with the mean value from all studies or with values from studies with the most similar conditions (i.e., length of follow up, background therapy) in the case of HRQoL score standard deviations. Missing values were treated in the same way as they were treated in the original studies.

## Results

After a search of MEDLINE, EMBASE, and Cochrane databases and the initial removal of duplicates we identified 1131 studies. After title and abstract screening 1058 studies were excluded (**Figure 1**), the full text of 73 studies were screened and 41 were included in the analysis.^28–37,43–72^ These studies had a median of 276 (interquartile range (IQR) 105, 464) participants and together analyzed 41,145 patients who were mostly male (76.5%) with a mean LVEF of 28% and a median follow up time of 5 months (IQR 3,8). Of the 41 included studies, 36 were multi-center, 16 were multinational, and 20 used the MLHFQ and 21 used the KCCQ. Ten studies evaluated the impact of beta-blockers on HRQoL, ten studies evaluated the impact of SGLT2i, 5 studies evaluated ACEi compared to ARNi, 3 studies each evaluated ivabradine and OM, 2 studies each evaluated ACEi and ARB, 1 study each evaluated H-ISDN, vericiguat, digoxin, MRA, ARB compared to ACEi, and ARB compared to ARNi (**Table 1**).

**Figure 1:**
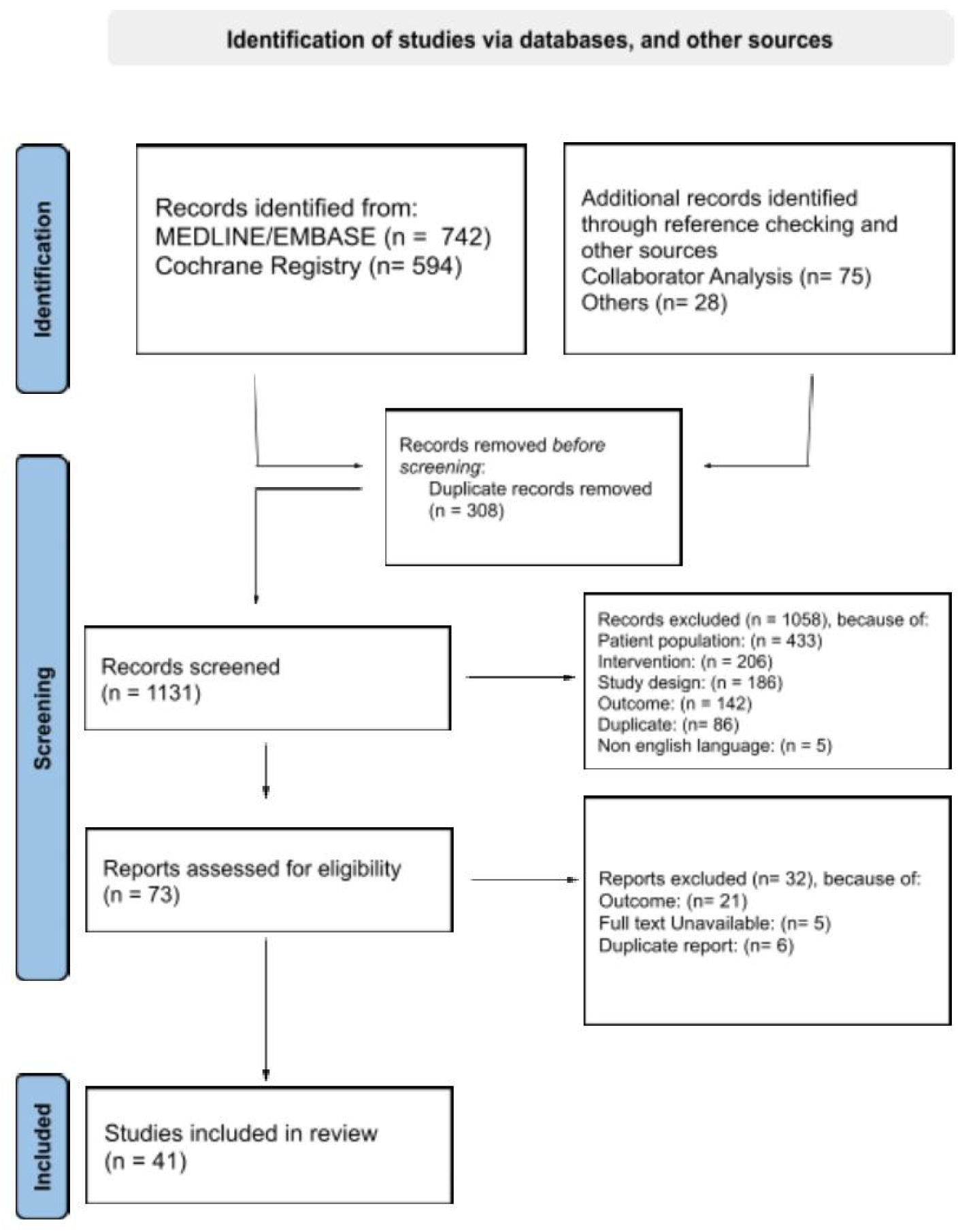
PRISMA flowchart.

**Table 1.** Study characteristics and outcomes by treatment arm.

|  | Treatment | Follow up, months | N randomized | Mean MLHFQ score change | Mean KCCQ overall summary score (OSS) change | Mean KCCQ total symptom score (TSS) change | Mean KCCQ clinical summary score (CSS) change | Number of participants discontinuing the drug in the group |
| --- | --- | --- | --- | --- | --- | --- | --- | --- |
| Pollock et al., 1990 | DIG | 3 | 7 | -6 | 4.11 | NR | NR | 4 |
|  | BB+DIG | 3 | 12 | -21 | 24.13 | NR | NR | 2 |
| Widimsky et al., 1995 | DIG | 3 | 48 | -8 | 6.78 | NR | NR | 6 |
|  | ACEi+DIG | 3 | 48 | -13 | 13.45 | NR | NR | 4 |
|  | ACEi+DIG | 3 | 53 | -8 | 6.78 | NR | NR | 2 |
|  | ACEi+DIG | 3 | 51 | -6 | 4.11 | NR | NR | 4 |
|  | ACEi+DIG | 3 | 48 | -9 | 8.11 | NR | NR | 6 |
| Bristow et al., 1996 | ACEi+DIG | 6 | 84 | -7.3 | 5.84 | NR | NR | 11 |
|  | ACEi+BB+DIG | 6 | 83 | -7.9 | 6.64 | NR | NR | 3 |
|  | ACEi+BB+DIG | 6 | 89 | -7.3 | 5.84 | NR | NR | 10 |
|  | ACEi+BB+DIG | 6 | 89 | -5.5 | 3.44 | NR | NR | 5 |
| Packer et al., 1996 | ACEi+DIG | 6 | 145 | -3.7 | 1.04 | NR | NR | 30 |
|  | ACEi+BB+DIG | 6 | 133 | -5.5 | 3.44 | NR | NR | 19 |
| Colucci et al., 1996 | ACEi+DIG | 12 | 134 | -4.9 | 2.64 | NR | NR | 14 |
|  | ACEi+BB+DIG | 12 | 232 | -2.4 | -0.70 | NR | NR | 17 |
| Cohn et al., 1997 | ACEi+DIG | 3 | 35 | -8.8 | 7.84 | NR | NR | 4 |
|  | ACEi+BB+DIG | 3 | 70 | -11.6 | 11.58 | NR | NR | 8 |
| Goldstein et al., 1999 | ACEi+DIG | 6 | 19 | -4.84 | 2.56 | NR | NR | 3 |
|  | ACEi+BB+DIG | 6 | 42 | -8.87 | 7.94 | NR | NR | 9 |
| Hjalmarson et al., 2000 | ACEi+DIG | 1 | 339 | 0.2 | -4.17 | NR | NR | NR |
|  | ACEi+BB+DIG | 12 | 331 | -0.7 | -2.97 | NR | NR | NR |
| Granger et al., 2000 | DIG | 3 | 91 | -4 | 1.44 | NR | NR | 12 |
|  | ARB+DIG | 3 | 179 | 0 | -3.90 | NR | NR | 31 |
| Beanlands et al., 2000 | Placebo | 3 | 21 | 1 | -5.24 | NR | NR | 1 |
|  | BB | 3 | 19 | 4 | -9.24 | NR | NR | 1 |
| De Milliano et al., 2001 | ACEi | 6 | 11 | -4.1 | 1.57 | NR | NR | 1 |
|  | ACEi+BB | 6 | 43 | -6.8 | 5.17 | NR | NR | 4 |
| Hutcheon et al., 2002 | Placebo | 2.5 | 37 | -5.4 | 3.31 | NR | NR | 2 |
|  | ACEi | 2.5 | 36 | -4.8 | 2.50 | NR | NR | 5 |
| Willenheimer et al., 2002 | ARB+BB | 3 | 70 | -0.7 | -2.97 | NR | NR | 6 |
|  | ACEi+BB | 3 | 71 | -0.9 | -2.70 | NR | NR | 14 |
| Lader et al., 2003 | ACEi | 12 | 291 | -5 | 2.77 | NR | NR | NR |
|  | ACEi+DIG | 12 | 298 | -1 | -2.57 | NR | NR | NR |
| Taylor et al., 2004 | ACEi+BB+DIG | 10 | 532 | -2.7 | -0.30 | NR | NR | NR |
|  | ACEi+BB+DIG+H-ISDN | 10 | 518 | -5.6 | 3.57 | NR | NR | NR |
| Majani et al., 2004 | ACEi+DIG | 23 | 1506 | 2.4 | -7.11 | NR | NR | NR |
|  | ACEi+ARB+DIG | 23 | 1504 | 0.4 | -4.44 | NR | NR | NR |
| Edes et al., 2005 | ACEi+DIG | 12 | 126 | -10.78 | 10.49 | NR | NR | 0 |
|  | ACEi+BB+DIG | 12 | 134 | -9.23 | 8.42 | NR | NR | 0 |
| Chan et al., 2007 | ARB+BB | 12 | 25 | -10.96 | 10.73 | NR | NR | 1 |
|  | ARB+BB+MRA | 12 | 23 | -12.3 | 12.52 | NR | NR | 2 |
| Ekman et al., 2011 | ACEi+BB+MRA | 12 | 976 | NR | 4.3 | 3.6 | 3.3 | NR |
|  | ACEi+BB+MRA+Ivabradine | 12 | 968 | NR | 6.7 | 4.6 | 5 | NR |
| Abdel-Salam et al., 2015 | ACEi+BB+MRA | 3 | 23 | -8.6 | 7.58 | NR | NR | 0 |
|  | ACEi+BB+MRA+Ivabradine | 3 | 20 | -12.4 | 12.65 | NR | NR | 0 |
| Lewis et al., 2017 | ACEi+BB+MRA | 8 | 3826 | NR | -0.14 | -0.61 | -0.29 | NR |
|  | ARNi+BB+MRA | 8 | 3797 | NR | 1.13 | 0.53 | 0.64 | NR |
| Nassif et al., 2019 | ACEi+BB+MRA | 3 | 132 | NR | 1.9 | -0.5 | 0.5 | 12 |
|  | ACEi+BB+MRA+SGLT2i | 3 | 131 | NR | 5.2 | 4.5 | 4.2 | 11 |
| Desai et al., 2019 | ACEi+BB | 3 | 233 | NR | 4.2 | NR | NR | 17 |
|  | ARNi+BB | 3 | 231 | NR | 8.7 | NR | NR | 16 |
| McMurray et al., 2019 | ACEi+BB+MRA | 8 | 2371 | NR | 4 | 3.3 | 3 | 249 |
|  | ACEi+BB+MRA+SGLT2i | 8 | 2373 | NR | 6.5 | 6.1 | 6 | 258 |
| Jensen et al., 2020 | ACEi+BB+MRA | 3 | 95 | NR | 1.9 | 1.1 | 0.5 | 1 |
|  | ACEi+BB+MRA+SGLT2i | 3 | 95 | NR | 2 | 3.2 | 3.3 | 0 |
| Felker et al., 2020 | ACEi+BB+MRA | 5 | 149 | NR | NR | 5 | 4.1 | 4 |
|  | ACEi+BB+MRA+OM | 5 | 149 | NR | NR | 9.9 | 7 | 12 |
|  | ACEi+BB+MRA+OM | 5 | 150 | NR | NR | 6.6 | 6.3 | 5 |
| Abraham et al., 2021 | ARNi+BB+MRA | 3 | 156 | NR | 6.4 | 3.65 | 4.82 | 13 |
|  | ARNi+BB+MRA+SGLT2i | 3 | 156 | NR | 9.65 | 7.29 | 8.2 | 15 |
| Santos-Gallego et al., 2021 | ARNi+BB | 6 | 42 | NR | 1.9 | NR | NR | 2 |
|  | ARNi+BB+SGLT2i | 6 | 42 | NR | 21 | NR | NR | 2 |
| Tsutsui et al., 2021 | ACEi+BB+M RA | 6 | 113 | NR | NR | NR | -3.49 | 13 |
|  | ARNi+BB+M RA | 6 | 112 | NR | NR | NR | -2.22 | 11 |
| Khandwalla et al., 2020 | ACEi+BB+M RA | 2 | 70 | NR | 4.19 | NR | NR | 8 |
|  | ARNi+BB+M RA | 2 | 70 | NR | 2.89 | NR | NR | 5 |
| Lee et al., 2021 | ACEi+BB+M RA | 9 | 53 | NR | NR | 4.2 | NR | 0 |
|  | ACEi+BB+M RA+SGLT2i | 9 | 52 | NR | NR | 0.7 | NR | 5 |
| Teerlink et al., 2021 | ACEi+BB+M RA | 6 | 3072 | NR | NR | 6.3 | NR | 50 |
|  | ACEi+BB+M RA+OM | 6 | 3076 | NR | NR | 5.8 | NR | 41 |
| Butler et al., 2021 | ACEi+BB+M RA | 12 | 1867 | NR | 5 | 5 | 4 | 335 |
|  | ACEi+BB+M RA+SGLT2i | 12 | 1863 | NR | 6.5 | 6.75 | 5.5 | 303 |
| Halle et al., 2021 | ACEi+BB+M RA | 3 | 98 | NR | 5.84 | 4.75 | 4.92 | 7 |
|  | ARNi+BB+M RA | 3 | 103 | NR | 8.11 | 8.29 | 7.33 | 4 |
| Mann et al., 2021 | ARB+BB+M RA | 6 | 168 | NR | 10.8 | NR | NR | 36 |
|  | ARNi+BB+M RA | 6 | 167 | NR | 11.8 | NR | NR | 49 |
| Ye et al., 2022 | ACEi+BB+M RA | 8 | 172 | NR | 5 | NR | 3 | 13 |
|  | ACEi+BB+M RA+Ivabradine | 8 | 170 | NR | 9 | NR | 7 | 13 |
| Palau et al., 2022 | ARNi+BB+M RA | 3 | 45 | -2.6 | -0.43 | NR | NR | 0 |
|  | ARNi+BB+M RA+SGLT2i | 3 | 45 | -6 | 4.11 | NR | NR | 0 |
| Spertus et al., 2022 | Placebo | 3 | 91 | NR | 6.6 | 4.2 | 3.9 | 18 |
|  | SGLT2i | 3 | 90 | NR | 9.8 | 9.1 | 7.5 | 13 |
| Butler et al., 2022 | ACEi+BB+MRA | 4 | 2524 | NR | 6.8 | 7.3 | 6.3 | 565 |
|  | ACEi+BB+MRA+Vericiguat | 4 | 2526 | NR | 7.8 | 6.3 | 6.3 | 610 |
| Lewis et al., 2022 | ARNi+BB+MRA | 5 | 91 | NR | NR | 1.8 | NR | 6 |
|  | ARNi+BB+MRA+OM | 5 | 185 | NR | NR | 0.3 | NR | 21 |
| McMurray et al., 2024 | ACEi+BB+MRA | 4 | 157 | NR | NR | 3.1 | NR | 12 |
|  | ACEi+BB+MRA+SGLT2i | 4 | 156 | NR | NR | 6.2 | NR | 9 |

Abbreviations: ACEI, angiotensin converting enzyme inhibitor; ARB, angiotensin-receptor blocker; ARNI, angiotensin receptor neprilysin inhibitor; BB, beta-blocker; DIG, digoxin; H-ISDN, hydralazine–isosorbide dinitrate; MRA, mineralocorticoid receptor antagonist; NR, not recorded; N, number.

### Risk of Bias and Certainty of Evidence

The within-study risk of bias was low as all studies were randomized, double-blind, placebo-controlled studies, most of which were multi-center. Two studies had major concerns of bias, as defined by the RoB2 tool, due to an imbalance in the number of missing patients between treatment groups without reported evidence that the outcomes of the missing participants were accounted for with an appropriate method.^55,72^ However, as one of the studies did not report the OSS change of their treatment groups, this study was not included in the main analysis.^72^ The other study with major concerns of bias, Willenheimer et al., was included in the main analysis, but sensitivity analyses showed that its removal did not significantly impact the main results (**Supplemental Appendix**).^55^ There was no evidence of funnel plot asymmetry or small study effects for the outcomes of change in HRQoL (Egger test p=0.818) or discontinuation (Egger test p=0.429) (**Supplemental Appendix**).

As the exclusion and inclusion criteria were very selective, indirectness was judged as low-very low. The minimally important clinical difference represents the minimum change which represents a noticeable improvement for a patient. In the KCCQ scale, a small clinical benefit has been defined as a change of 5 units. Consequently, the effect size to evaluate the imprecision and heterogeneity in the study was defined as 5 units of score change in our study.^73^ Due to the high variability in HRQoL scores observed across all the studies, there was a high level of imprecision in many treatment comparisons. Overall heterogeneity for the additive network meta-analysis model was high (I^2^ 61.9%) therefore the results from the random effects model were considered the primary results. Incoherence was not significant and the confidence rating was high or moderate for all clinically relevant treatment comparisons. The confidence in the results of comparisons between treatments was generally lower for many of the comparisons such as which relied on indirect evidence from a large trial and a smaller trial, such as ACEi compared with ARNi + BB + MRA + SGLT2i or ARB + BB compared with SGLT2i, and those that included data from the Willenheimer et al. trial.^55^

### Component Analysis

Out of the 41 studies included in the NMA only 35 reported either the KCCQ OSS or the MLHFQ score;^6,28–37,43–72^ these studies were included in the main analysis of change in HRQoL measured through the change in OSS and converted MLHFQ scores (**Figure 2**). It was not possible to analyze the other domains of the KCCQ as there were not enough studies to build an adequate network; 14, 15, and 9 studies reported the clinical summary score domain, the total symptom score domain, and the physical limitation score domain in sufficient detail for analysis respectively. The main analysis indicated that the therapy with the largest effect on HRQoL was H-ISDN [MD 3.87 (95% CI, -0.73 to 8.47)], however, this effect was not statistically significant. The treatment with the next largest effect was SGLT2i [MD 3.37 (95% CI, 1.44 to 5.30)] which led to the greatest improvement in QoL, followed by ivabradine [MD 3.26 (95% CI, 0.08 to 6.43)], then ARNi [MD 2.62 (95% CI, -3.24 to 8.47)] and then vericiguat [MD 1.00 (95% CI, -3.18 to 5.18)]. BB, ACEi, and ARB had a neutral impact on HRQoL, and digoxin improved HRQoL significantly less than placebo [MD -5.34 (95% CI, -10.30 to -0.38)] (**Figure 3**).

**Figure 2:**
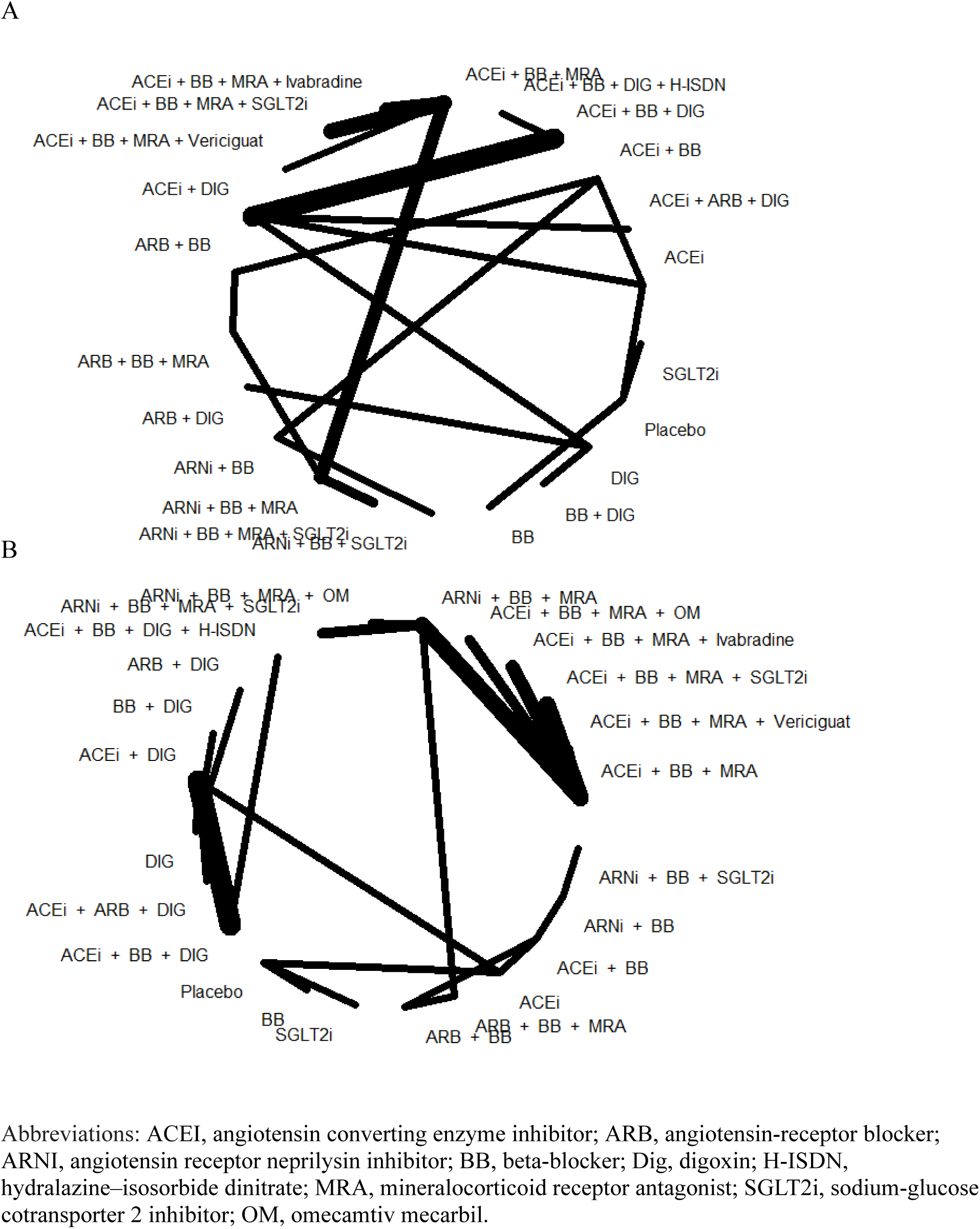
Network connection diagram for change in quality of life (A) and discontinuation (B) outcomes.

**Figure 3:**
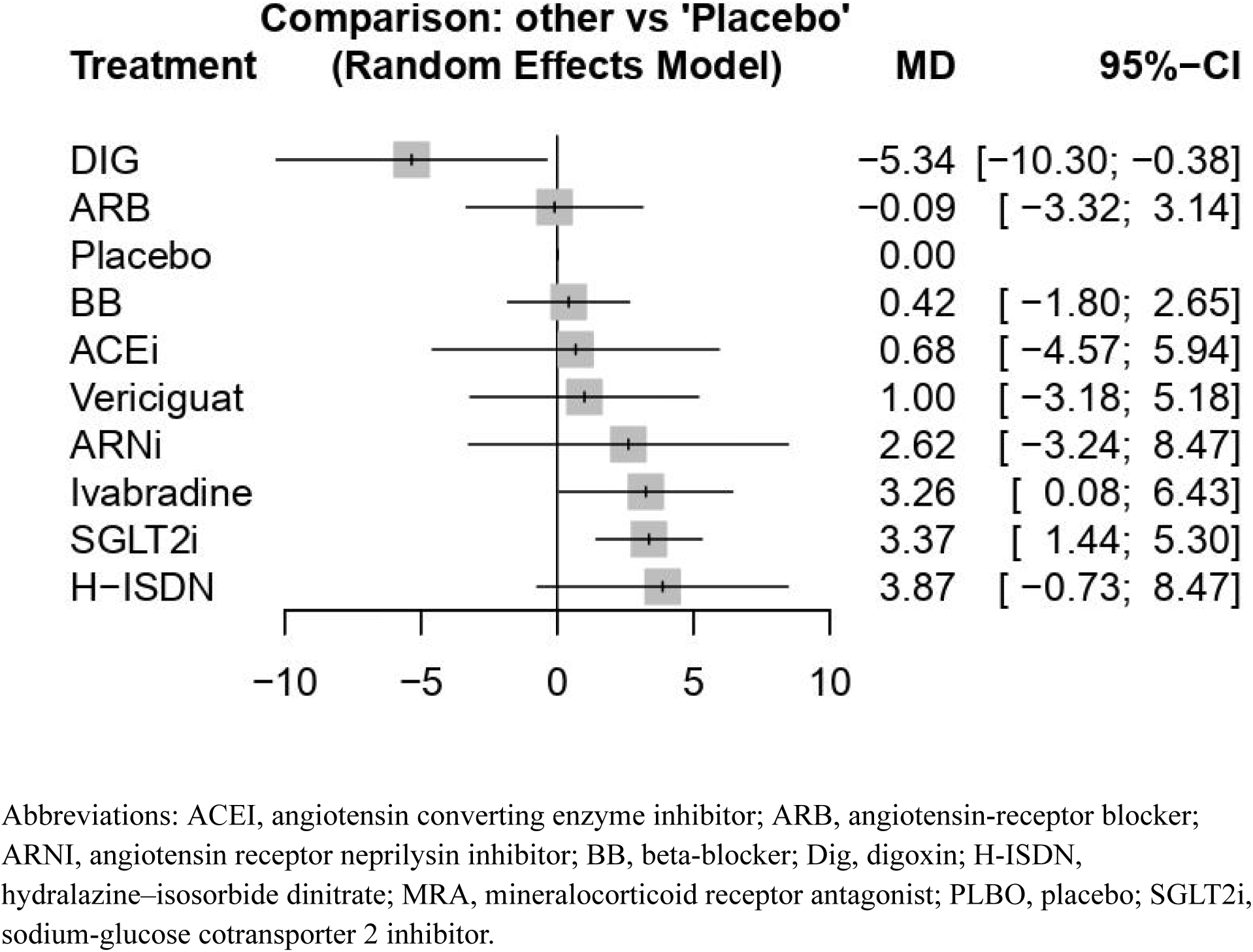
Forest plot showing the mean difference in HRQoL score change in individual treatments against placebo.

### Additive Network Analysis

A combination of ARNi + BB + MRA + SGLT2i [MD 7.11 (95% CI, -0.99 to 15.22)] had the largest effect on HRQoL, but this effect was not significant. The combination of ARNi + BB + SGLT2i [MD 5.33 (95% CI, 0.40 to 10.25)] had the largest statistically significant effect on quality of life. Other combinations such as ACEi + BB + MRA + SGLT2i [MD 5.32 (95% CI, -2.63 to 13.26)], ACEi + BB + MRA + ivabradine [MD 5.24 (95% CI, -3.07 to 13.55)], and ARNi + BB + MRA [MD 3.81 (95% CI, -4.09 to 11.71)] also did not have a significant effect (**Figure 4**). Other combinations of ACEi/ARB/ARNi with BB and MRA generally had an unclear impact on HRQoL and most combinations involving digoxin had a negative impact on HRQoL compared with placebo even when paired with ACEi/ARB or BB. The *P*-score values given in the **Supplemental Appendix** demonstrate a similar pattern of ranking. The combination of ARNi + BB + MRA + SGLT2i did not improve HRQoL significantly better than the combination of ACEi + BB + MRA + SGLT2i or the combination of ARNi + BB + SGLT2i, but was significantly more effective than a combination of ARNi + BB + MRA [MD 3.3 (95% CI, 1.5 to 5.11); p <0.001)] (**Supplemental Appendix**).

**Figure 4:**
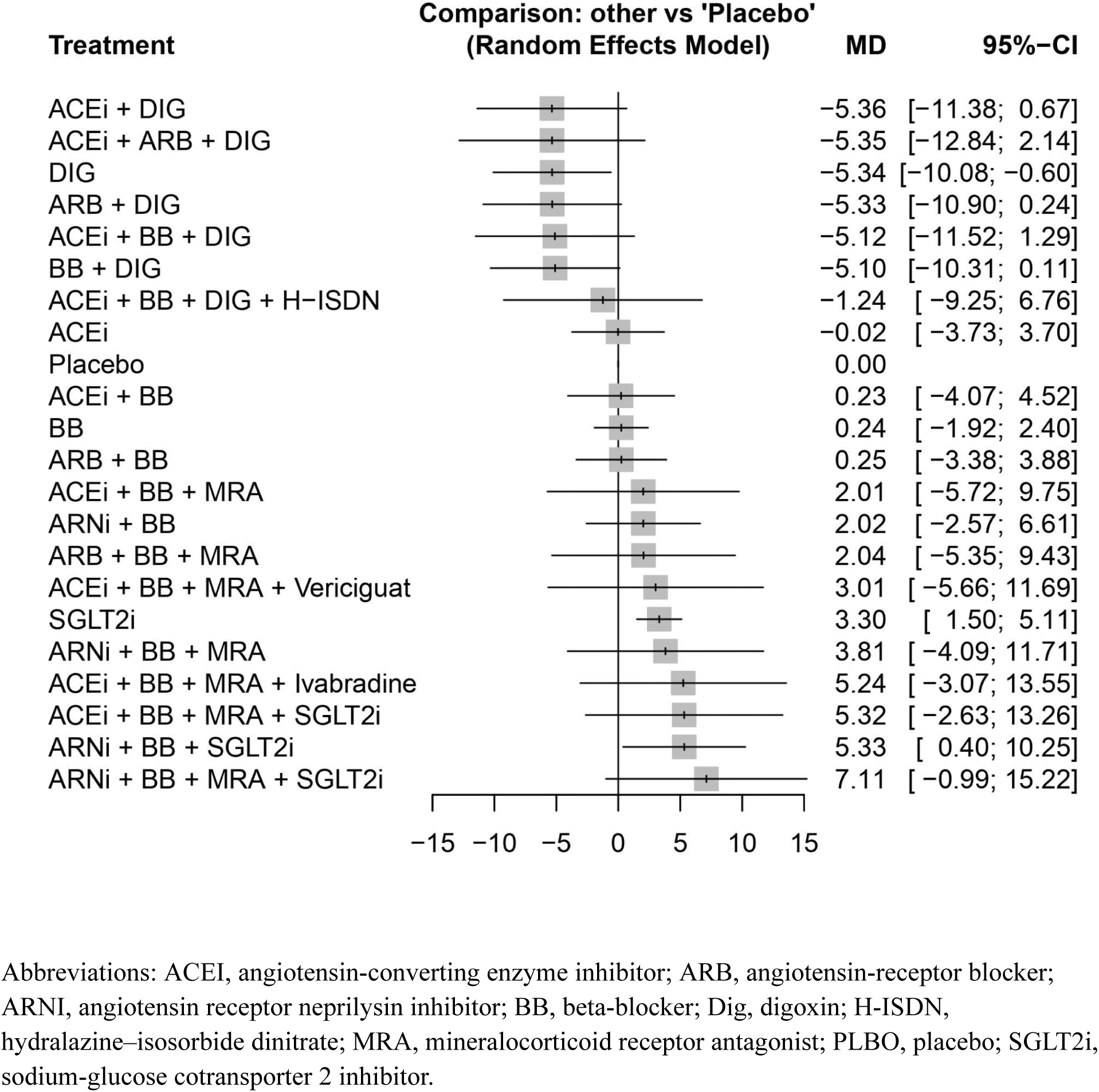
Forest plot showing the mean difference in quality of life score change in various treatment combinations against placebo.

The discontinuation of study interventions was recorded in 35 studies comprising 2977 discontinuations and there was low heterogeneity (I^2 19.2%) in the network. Compared to placebo, participants were significantly more likely to discontinue the use of ACEi [HR 2.61 (95% CI, 1.33 to 5.11)], and ARNi [HR 2.21 (95% CI, 1.11 to 4.41)], but not any of the other interventions (**Supplemental Appendix**).

### Sensitivity Analysis

Four sensitivity analyses were conducted to supplement the main results of this study. The first sensitivity analysis found that removing the studies that were deemed to have major concerns of bias from the primary analysis did not change the results (**Supplemental Appendix**). The second sensitivity analysis evaluated the sensitivity of the results to the conversion of MLHFQs score to the KCCQ score scale. This was achieved by comparing a subnetwork with only trials that have converted scores to a subnetwork of only trials with the original MLHFQ scores, and the result was largely the same (**Supplemental Appendix**). The third sensitivity analysis evaluated the validity of the consideration of the trials against a background of ARNi if more than 40% of the patients using ACEi/ARNi/ARB in either arm were on ARNi, or a background of ACEi if less than 40% of patients were on ARNi. We evaluated all of the trials with mixed ACEi/ARNi/ARB use on a background of ACEi instead of ARNi and the results remained largely the same (**Supplemental Appendix**). The fourth sensitivity analysis found that removing the studies that did not describe the background therapy did not change the results (**Supplemental Appendix**).

## Discussion

This study evaluated the effect of interventions efficacious in HFrEF on HRQoL. Our study has two major findings. First, evidence with high confidence showed that a combination of ARNI + BB + MRA + SGLT2i had the greatest impact on HRQoL, followed by combinations of ARNi + BB + SGLT2i or ACEi + BB + MRA + SGLT2i. This complements the findings of a prior NMA focused on other clinical outcomes, and supports the guideline statements surrounding the use of all 4 medication classes concurrently.^4,5,21,74^ Second, the addition of SGLT2i and ivabradine to other treatment combinations improved HRQoL scores while the addition of digoxin to the treatment improved HRQoL less than placebo. Clinicians should consider adding medications that improve HRQoL and discontinuing medications without strong evidence of benefit.

The current GDMT for HFrEF involves the initiation and up titration of a combination of ARNi + BB + MRA + SGLT2i if possible as it is the most effective for improving survival.

However, our study results could not confirm this combination was the most effective for improving HRQoL statistically, but it was favored numerically with a mean difference of 7.1 points. We also could not confirm whether the replacement of SGLT2i with vericiguat or ivabradine in combination with ACEi, BB, and MRA resulted in a change in HRQoL as no significant difference in effect was found. The lack of significance for many results may not be an accurate reflection of the true effect of the treatment combinations and instead it may be due partially to the variability of the measure of change in HRQoL score and the lack of direct evidence for many of the comparisons. To our knowledge this is the first study to build a network meta-analysis for the evaluation of the efficacy of HFrEF interventions in improving HRQoL, however, similar analyses evaluating these treatment combinations for their impact on mortality and hospitalization, which are outcomes that can influence HRQoL, have found similar rankings of treatment combinations, but were often able to identify significant treatment effects.^75–77^ Thus, while more direct comparisons and higher certainty evidence are needed, the current evidence together with the results of similar studies supports the immediate initiation and uptitration of a quadruple therapy consisting of ARNi + BB + MRA, and SGLT2i or a triple therapy of ARNI + BB + SGLT2i for the majority of patients with HFrEF. More evidence is also needed to determine if vericiguat or ivabradine can or should be added to quadruple therapy or used in combination with some parts of GDMT to increase HRQoL for any subgroup of patients with HFrEF.

Concerning the effect of each treatment component individually, our results generally agree with previously conducted non-network based meta analyses. Results from recently conducted meta analyses support our conclusions concerning the efficacy of SGLT2i and ivabradine in improving HRQoL as it has been demonstrated previously that SGLT2i and ivabradine can significantly improve HRQoL.^6,18,19^ Also, similar to the findings of past research, BB and ACEi did not significantly impact on HRQoL.^6^ However, our study suggests that digoxin, which was previously shown to be neutral in its impact on HRQoL, may not significantly improve HRQoL more than placebo.^6^ Further, ARNi, ARB, and H-ISDN, which have been suggested to significantly improve HRQoL did not have a statistically significant impact on HRQoL in our study.^6–8^ The impact of vericiguat on HRQoL has not been evaluated previously, and in our study it did not have a significant impact compared to placebo. We were unable to assess the efficacy of omecamtiv mecarbil and MRA due to the lack of trials which evaluated the effect of the interventions on the KCCQ OSS. Our results suggest that the addition of ivabradine could be considered after GDMT is optimized, whereas more evidence may be needed to determine the efficacy of vericiguat and H-ISDN in improving HRQoL before it is considered for addition to GDMT, particularly since only one trial evaluated the impact of H-ISDN and this trial only involved patients who self-identified as Black.^57^ Digoxin, however, should not be considered for addition to GDMT as it may result in a clinically meaningful reduction in HRQoL improvement. This is likely to be the case because particularly before the use of multiple imputation methods was computationally accessible, HRQoL measurement methods heavily penalized treatment groups for patient deaths by imputing the worst score possible for missing data.^78,79^ The Digitalis Investigation Group (DIG) trial - which was the only trial which evaluated digoxin directly in our study - used the worst score possible imputation method as a sensitivity analysis and patient scores using this method were found to be similar to the scores used in the main analysis which suggests that the imputed numbers may have been lower than the true scores of participants.^80^ As digoxin may not reduce mortality in patients with HFrEF the potential toxicity and risks of digoxin treatment may not be compensated in the way that other medications such as ACEi and ARNi are, due to their beneficial effects on mortality.^21,80,81^ Consequently, digoxin appears to improve HRQoL less than placebo in our study findings.

The conversion between the outcome measure of the study and the KCCQ OSS is direct so that the estimated MD of any treatment combination also describes the mean change in the KCCQ OSS which may be experienced by a patient who is initiated on this treatment. This means that patients who are initiated on a combination of ARNi + BB + MRA + SGLT2i may on average experience a small-medium clinically-meaningful HRQoL improvement and for combinations of ARNi + BB + SGLT2i, ACEi + BB + MRA + SGLT2i, or ACEi + BB + MRA + ivabradine a small clinically meaningful HRQoL improvement may be experienced on average. While many of the treatment combinations did not significantly improve HRQoL due to the highly variable nature of the HRQoL score, the evidence is still applicable to a significant portion of the population of patients with HFrEF and can be an important consideration when making treatment decisions. Additionally, due to the variable nature of the experience of HRQoL, the mean change has been suggested to be an incomplete description of the change in HRQoL that a patient may experience. Patients are not likely to experience a mean improvement in HRQoL but rather a deterioration, no improvement or an improvement in HRQoL; consequently, it is suggested to summarize the intervention’s impact on HRQoL by considering and comparing the proportion of patients who experience a change of 5 units, 10 units and 20 units in the KCCQ - which have been defined by Spertus et al. as small, medium and large clinical changes respectively - in either direction in addition to the mean change in HRQoL due to the intervention.^73^ Thus, the evaluated treatment combinations may result in a more significant benefit than estimated for a smaller subset of the population which uses them. Trials have not traditionally reported HRQoL changes in this way, so it was not possible to use this measurement for our study. However, it should be considered for future research evaluating the impact of HF treatments on HRQoL.

## Strengths and Limitations

Several strengths and limitations should be considered. First, while this is the largest and only study to evaluate the impact of combinations of treatments for heart failure with reduced ejection fraction on HRQoL using a network meta analysis and systematic review methodology it is reliant on studies conducted over 4 decades and may be susceptible to secular changes in care.

Nevertheless, the inclusion of 41 placebo or active controlled, randomized, double blind trials, the majority of which are multicenter trials represents the best available evidence for the impact of efficacious HFrEF treatments on patients’ HRQoL. Second, bias is commonly identified in meta-analyses, however, the main analysis for the most clinically relevant treatment combinations largely relied on high confidence evidence with little detected bias. Third, two different HRQoL instruments were used and conversion can be challenging. Therefore, we conducted sensitivity analyses which showed that the conversion of scores from one score scale to the other and the bias that was detected in some studies did not influence the results of our study. However, many treatment combinations in our study did not have a statistically significant impact on HRQoL possibly due to the natural variability in HRQoL experiences within study treatment groups and the lack of direct evidence for comparisons. Fourth, while the heterogeneity of the standard NMA model was low, the additive NMA model showed high heterogeneity which was partially compensated for by using a random effects model. Fifth, we recognize that due to the additive nature of medication prescription in heart failure, comparing the efficacy of medications which are frequently used together has not occurred. As a result, our study findings rely largely on indirect comparisons of treatments, however, while more direct data may strengthen our results, reliance on indirect comparisons of evidence from the best currently available subset of trials does not reduce the significance of the insights or the alignment of the methodology with the topic and objectives of this study. Finally, we were unable to account for the dosages of study treatments or the variability in length of follow up between studies.

## Conclusion

The results of this study indicate the most effective treatment combinations and additional therapies for improving HRQoL. This provides clinically meaningful information that can help patients and clinicians achieve patient goals more effectively.

## Data Availability

All data produced in the present study are available upon reasonable request to the authors.

## Acknowledgements

The authors would like to gratefully acknowledge Lisa Soulard for editorial assistance in preparing the manuscript.

## Supplemental Material

Tables S1–S4

Figure S1-S7

Search Terms

Protocol

## References

1. Yan T, Zhu S, Yin X, Xie C, Xue J, Zhu M, Weng F, Zhu S, Xiang B, Zhou X, et al. Burden, Trends, and Inequalities of Heart Failure Globally, 1990 to 2019: A Secondary Analysis Based on the Global Burden of Disease 2019 Study. J. Am. Heart Assoc. 2023;12:e027852.

2. GBD 2017 Disease and Injury Incidence and Prevalence Collaborators. Global, regional, and national incidence, prevalence, and years lived with disability for 354 diseases and injuries for 195 countries and territories, 1990-2017: a systematic analysis for the Global Burden of Disease Study 2017. Lancet. 2018;392:1789–1858.

3. Tran DT, Ohinmaa A, Thanh NX, Howlett JG, Ezekowitz JA, McAlister FA, Kaul P. The current and future financial burden of hospital admissions for heart failure in Canada: a cost analysis. CMAJ Open. 2016;4:E365–E370.

4. Heidenreich PA, Bozkurt B, Aguilar D, Allen LA, Byun JJ, Colvin MM, Deswal A, Drazner MH, Dunlay SM, Evers LR, et al. 2022 AHA/ACC/HFSA Guideline for the Management of Heart Failure: A Report of the American College of Cardiology/American Heart Association Joint Committee on Clinical Practice Guidelines. Circulation. 2022;145:e895–e1032.

5. McDonagh TA, Metra M, Adamo M, Gardner RS, Baumbach A, Böhm M, Burri H, Butler J, Čelutkienė J, Chioncel O, et al. 2021 ESC Guidelines for the diagnosis and treatment of acute and chronic heart failure: Developed by the Task Force for the diagnosis and treatment of acute and chronic heart failure of the European Society of Cardiology (ESC) With the special contribution of the Heart Failure Association (HFA) of the ESC. Eur Heart J . 2022;42:3599–3726.

6. Turgeon RD, Barry AR, Hawkins NM, Ellis UM. Pharmacotherapy for heart failure with reduced ejection fraction and health-related quality of life: a systematic review and meta-analysis. Eur. J. Heart Fail. 2021;23:578–589.

7. Shah YR, Turgeon RD. Impact of SGLT2 Inhibitors on Quality of Life in Heart Failure Across the Ejection Fraction Spectrum: Systematic Review and Meta-analysis. CJC Open. 2024;6:639–648.

8. Song Y, Zhao Z, Zhang J, Zhao F, Jin P. Effects of sacubitril/valsartan on life quality in chronic heart failure: A systematic review and meta-analysis of randomized controlled trials. Front Cardiovasc Med. 2022;9:922721.

9. Yang H-R, Xu X-D, Shaikh AS, Zhou B-T. Efficacy and Safety of Sacubitril/Valsartan Compared With ACEI/ARB on Health-Related Quality of Life in Heart Failure Patients: A Meta-Analysis. Ann. Pharmacother. 2023;57:907–917.

10. Ibrahim R, Olagunju A, Terrani K, Takamatsu C, Khludenev G, William P. KCCQ total symptom score, clinical outcome measures, and adverse events associated with omecamtiv mecarbil for heart failure with reduced ejection fraction: a systematic review and meta-analysis of randomized controlled trials. Clin. Res. Cardiol. 2023;112:1067–1076.

11. Aimo A, Pateras K, Stamatelopoulos K, Bayes-Genis A, Lombardi CM, Passino C, Emdin M, Georgiopoulos G. Relative Efficacy of Sacubitril-Valsartan, Vericiguat, and SGLT2 Inhibitors in Heart Failure with Reduced Ejection Fraction: a Systematic Review and Network Meta-Analysis. Cardiovasc. Drugs Ther. 2021;35:1067–1076.

12. Patoulias D, Papadopoulos C, Kalogirou M-S, Katsimardou A, Toumpourleka M, Doumas M. Updated Meta-analysis Assessing the Effect of Sodium-Glucose Co-transporter-2 Inhibitors on Surrogate End points in Patients With Heart Failure With Reduced Ejection Fraction. Am. J. Cardiol. 2020;137:130–132.

13. Butler J, Usman MS, Khan MS, Greene SJ, Friede T, Vaduganathan M, Filippatos G, Coats AJS, Anker SD. Efficacy and safety of SGLT2 inhibitors in heart failure: systematic review and meta-analysis. ESC Heart Fail. 2020;7:3298–3309.

14. Chambergo-Michilot D, Tauma-Arrué A, Loli-Guevara S. Effects and safety of SGLT2 inhibitors compared to placebo in patients with heart failure: A systematic review and meta-analysis. Int J Cardiol Heart Vasc. 2021;32:100690.

15. Kumar K, Kheiri B, Simpson TF, Osman M, Rahmouni H. Sodium-Glucose Cotransporter-2 Inhibitors in Heart Failure: A Meta-Analysis of Randomized Clinical Trials. Am. J. Med. 2020;133:e625–e630.

16. Salah HM, Al’Aref SJ, Khan MS, Al-Hawwas M, Vallurupalli S, Mehta JL, Mounsey JP, Greene SJ, McGuire DK, Lopes RD, et al. Effect of sodium-glucose cotransporter 2 inhibitors on cardiovascular and kidney outcomes-Systematic review and meta-analysis of randomized placebo-controlled trials. Am. Heart J. 2021;232:10–22.

17. Zheng X-D, Qu Q, Jiang X-Y, Wang Z-Y, Tang C, Sun J-Y. Effects of Dapagliflozin on Cardiovascular Events, Death, and Safety Outcomes in Patients with Heart Failure: A Meta-Analysis. Am. J. Cardiovasc. Drugs. 2021;21:321–330.

18. He Z, Yang L, Nie Y, Wang Y, Wang Y, Niu X, Bai M, Yao Y, Zhang Z. Effects of SGLT-2 inhibitors on health-related quality of life and exercise capacity in heart failure patients with reduced ejection fraction: A systematic review and meta-analysis. Int J.Cardiol. 2021;345:83–88.

19. Usman MS, Hamid A, Siddiqi TJ, Shurjeel Q, Qazi S, Butler J. Effect of SGLT2 Inhibitors on Health-Related Quality of Life in Patients with Heart Failure: A Systematic Review and Meta-Analysis. J. Am. Coll. Cardiol. 2023;81:756.

20. Sephien A, Ghobrial M, Reljic T, Prida X, Nerella N, Kumar A. Efficacy of SGLT2 inhibitors in patients with heart failure: An overview of systematic reviews. Int. J. Cardiol. 2023;377:79–85.

21. Tromp J, Ouwerkerk W, van Veldhuisen DJ, Hillege HL, Richards AM, van der Meer P, Anand IS, Lam CSP, Voors AA. A Systematic Review and Network Meta-Analysis of Pharmacological Treatment of Heart Failure With Reduced Ejection Fraction. JACC Heart Fail. 2022;10:73–84.

22. Kraai IH, Vermeulen KM, Luttik MLA, Hoekstra T, Jaarsma T, Hillege HL. Preferences of heart failure patients in daily clinical practice: quality of life or longevity? Eur. J. Heart Fail. 2013;15:1113–1121.

23. Albarrati AM, Altimani R, Almogbel O, Alnahdi AH, Almurdi MM, Abuammah A, Nazer R. Reliability and Validity of Kansas City Cardiomyopathy Questionnaire in Arabic Patients with Chronic Heart Failure. Medicina. 2023;59:1910.

24. Bilbao A, Escobar A, García-Perez L, Navarro G, Quirós R. The Minnesota living with heart failure questionnaire: comparison of different factor structures. Health Qual. Life Outcomes. 2016;14:23.

25. Yee D, Novak E, Platts A, Nassif ME, LaRue SJ, Vader JM. Comparison of the Kansas City Cardiomyopathy Questionnaire and Minnesota Living With Heart Failure Questionnaire in Predicting Heart Failure Outcomes. Am. J. Cardiol. 2019;123:807–812.

26. Nassif ME, Tang Y, Cleland JG, Abraham WT, Linde C, Gold MR, Young JB, Daubert JC, Sherfesee L, Schaber D, et al. Precision Medicine for Cardiac Resynchronization. Circ. Heart Fail. 2017;10:e004111.

27. Greene SJ, Fonarow GC, DeVore AD, Sharma PP, Vaduganathan M, Albert NM, Duffy CI, Hill CL, McCague K, Patterson JH, et al. Titration of Medical Therapy for Heart Failure With Reduced Ejection Fraction. J. Am. Coll. Cardiol. 2019;73:2365–2383.

28. Lewis GD, Voors AA, Cohen-Solal A, Metra M, Whellan DJ, Ezekowitz JA, Böhm M, Teerlink JR, Docherty KF, Lopes RD, et al. Effect of Omecamtiv Mecarbil on Exercise Capacity in Chronic Heart Failure With Reduced Ejection Fraction: The METEORIC-HF Randomized Clinical Trial. JAMA. 2022;328:259–269.

29. Palau P, Amiguet M, Domínguez E, Sastre C, Mollar A, Seller J, Garcia Pinilla JM, Larumbe A, Valle A, Gómez Doblas JJ, et al. Short-term effects of dapagliflozin on maximal functional capacity in heart failure with reduced ejection fraction (DAPA-VO2 ): a randomized clinical trial. Eur. J. Heart Fail. 2022;24:1816–1826.

30. Spertus JA, Birmingham MC, Nassif M, Damaraju CV, Abbate A, Butler J, Lanfear DE, Lingvay I, Kosiborod MN, Januzzi JL. The SGLT2 inhibitor canagliflozin in heart failure: the CHIEF-HF remote, patient-centered randomized trial. Nat. Med. 2022;28:809–813.

31. Butler J, Stebbins A, Melenovský V, Sweitzer NK, Cowie MR, Stehlik J, Khan MS, Blaustein RO, Ezekowitz JA, Hernandez AF, et al. Vericiguat and Health-Related Quality of Life in Patients With Heart Failure With Reduced Ejection Fraction: Insights From the VICTORIA Trial. Circ. Heart Fail. 2022;15:e009337.

32. Butler J, Anker SD, Filippatos G, Khan MS, Ferreira JP, Pocock SJ, Giannetti N, Januzzi JL, Piña IL, Lam CSP, et al. Empagliflozin and health-related quality of life outcomes in patients with heart failure with reduced ejection fraction: the EMPEROR-Reduced trial. Eur. Heart J. 2021;42:1203–1212.

33. Abraham WT, Lindenfeld J, Ponikowski P, Agostoni P, Butler J, Desai AS, Filippatos G, Gniot J, Fu M, Gullestad L, et al. Effect of empagliflozin on exercise ability and symptoms in heart failure patients with reduced and preserved ejection fraction, with and without type 2 diabetes. Eur. Heart J. 2021;42:700–710.

34. Santos-Gallego CG, Vargas-Delgado AP, Requena-Ibanez JA, Garcia-Ropero A, Mancini D, Pinney S, Macaluso F, Sartori S, Roque M, Sabatel-Perez F, et al. Randomized Trial of Empagliflozin in Nondiabetic Patients With Heart Failure and Reduced Ejection Fraction. J. Am. Coll. Cardiol. 2021;77:243–255.

35. Jensen J, Omar M, Kistorp C, Poulsen MK, Tuxen C, Gustafsson I, Køber L, Gustafsson F, Faber J, Fosbøl EL, et al. Twelve weeks of treatment with empagliflozin in patients with heart failure and reduced ejection fraction: A double-blinded, randomized, and placebo-controlled trial. Am. Heart J. 2020;228:47–56.

36. Nassif ME, Windsor SL, Tang F, Khariton Y, Husain M, Inzucchi SE, McGuire DK, Pitt B, Scirica BM, Austin B, et al. Dapagliflozin Effects on Biomarkers, Symptoms, and Functional Status in Patients With Heart Failure With Reduced Ejection Fraction: The DEFINE-HF Trial. Circulation. 2019;140:1463–1476.

37. Desai AS, Solomon SD, Shah AM, Claggett BL, Fang JC, Izzo J, McCague K, Abbas CA, Rocha R, Mitchell GF, et al. Effect of Sacubitril-Valsartan vs Enalapril on Aortic Stiffness in Patients With Heart Failure and Reduced Ejection Fraction: A Randomized Clinical Trial. JAMA. 2019;322:1077–1084.

38. Rücker G, Schwarzer G. Ranking treatments in frequentist network meta-analysis works without resampling methods. BMC Med. Res. Methodol. 2015;15:58.

39. Sterne JAC, Savović J, Page MJ, Elbers RG, Blencowe NS, Boutron I, Cates CJ, Cheng H-Y, Corbett MS, Eldridge SM, et al. RoB 2: a revised tool for assessing risk of bias in randomised trials. BMJ. 2019;366:l4898.

40. Nikolakopoulou A, Higgins JPT, Papakonstantinou T, Chaimani A, Del Giovane C, Egger M, Salanti G. CINeMA: An approach for assessing confidence in the results of a network meta-analysis. PLoS Med. 2020;17:e1003082.

41. Sterne JA, Egger M, Smith GD. Systematic reviews in health care: Investigating and dealing with publication and other biases in meta-analysis. BMJ. 2001;323:101–105.

42. Higgins JPT, Thompson SG. Quantifying heterogeneity in a meta-analysis. Stat. Med. 2002;21:1539–1558.

43. Pollock SG, Lystash J, Tedesco C, Craddock G, Smucker ML. Usefulness of bucindolol in congestive heart failure. Am. J. Cardiol. 1990;66:603–607.

44. Widimský J, Kremer HJ, Jerie P, Uhlír O. Czech and Slovak spirapril intervention study (CASSIS). A randomized, placebo and active-controlled, double-blind multicentre trial in patients with congestive heart failure. Eur. J. Clin. Pharmacol. 1995;49:95–102.

45. Bristow MR, Gilbert EM, Abraham WT, Adams KF, Fowler MB, Hershberger RE, Kubo SH, Narahara KA, Ingersoll H, Krueger S, et al. Carvedilol produces dose-related improvements in left ventricular function and survival in subjects with chronic heart failure. MOCHA Investigators. Circulation. 1996;94:2807–2816.

46. Packer M, Colucci WS, Sackner-Bernstein JD, Liang CS, Goldscher DA, Freeman I, Kukin ML, Kinhal V, Udelson JE, Klapholz M, et al. Double-blind, placebo-controlled study of the effects of carvedilol in patients with moderate to severe heart failure. The PRECISE Trial. Prospective Randomized Evaluation of Carvedilol on Symptoms and Exercise. Circulation. 1996;94:2793–2799.

47. Colucci WS, Packer M, Bristow MR, Gilbert EM, Cohn JN, Fowler MB, Krueger SK, Hershberger R, Uretsky BF, Bowers JA, et al. Carvedilol inhibits clinical progression in patients with mild symptoms of heart failure. US Carvedilol Heart Failure Study Group. Circulation. 1996;94:2800–2806.

48. Cohn JN, Fowler MB, Bristow MR, Colucci WS, Gilbert EM, Kinhal V, Krueger SK, Lejemtel T, Narahara KA, Packer M, et al. Safety and efficacy of carvedilol in severe heart failure. The U.S. Carvedilol Heart Failure Study Group. J. Card. Fail. 1997;3:173–179.

49. Goldstein S, Kennedy HL, Hall C, Anderson JL, Gheorghiade M, Gottlieb S, Jessup M, Karlsberg RP, Friday G, Haskell L. Metoprolol CR/XL in patients with heart failure: A pilot study examining the tolerability, safety, and effect on left ventricular ejection fraction. Am. Heart J. 1999;138:1158–1165.

50. Hjalmarson A, Goldstein S, Fagerberg B, Wedel H, Waagstein F, Kjekshus J, Wikstrand J, El Allaf D, Vítovec J, Aldershvile J, et al. Effects of controlled-release metoprolol on total mortality, hospitalizations, and well-being in patients with heart failure: the Metoprolol CR/XL Randomized Intervention Trial in congestive heart failure (MERIT-HF). MERIT-HF Study Group. JAMA. 2000;283:1295–1302.

51. Granger CB, Ertl G, Kuch J, Maggioni AP, McMurray J, Rouleau JL, Stevenson LW, Swedberg K, Young J, Yusuf S, et al. Randomized trial of candesartan cilexetil in the treatment of patients with congestive heart failure and a history of intolerance to angiotensin-converting enzyme inhibitors. Am. Heart J. 2000;139:609–617.

52. Beanlands RS, Nahmias C, Gordon E, Coates G, deKemp R, Firnau G, Fallen E. The effects of beta(1)-blockade on oxidative metabolism and the metabolic cost of ventricular work in patients with left ventricular dysfunction: A double-blind, placebo-controlled, positron-emission tomography study. Circulation. 2000;102:2070–2075.

53. de Milliano PAR, de Groot AC, Tijssen JGP, van Eck-Smit BLF, Van Zwieten PA, Lie KI. Beneficial effects of metoprolol on myocardial sympathetic function: Evidence from a randomized, placebo-controlled study in patients with congestive heart failure. Am. Heart J. 2002;144:E3.

54. Hutcheon SD, Gillespie ND, Crombie IK, Struthers AD, McMurdo MET. Perindopril improves six minute walking distance in older patients with left ventricular systolic dysfunction: a randomised double blind placebo controlled trial. Heart. 2002;88:373–377.

55. Willenheimer R, Helmers C, Pantev E, Rydberg E, Löfdahl P, Gordon A, Heart Failure Valsartan Exercise Capacity Evaluation Study Group. Safety and efficacy of valsartan versus enalapril in heart failure patients. Int. J. Cardiol. 2002;85:261–270.

56. Lader E, Egan D, Hunsberger S, Garg R, Czajkowski S, McSherry F. The effect of digoxin on the quality of life in patients with heart failure. J. Card. Fail. 2003;9:4–12.

57. Taylor Anne L., Ziesche Susan, Yancy Clyde, Carson Peter, D’Agostino Ralph, Ferdinand Keith, Taylor Malcolm, Adams Kirkwood, Sabolinski Michael, Worcel Manuel, et al. Combination of Isosorbide Dinitrate and Hydralazine in Blacks with Heart Failure. N. Engl. J. Med. 351:2049–2057.

58. Majani G, Giardini A, Opasich C, Glazer R, Hester A, Tognoni G, Cohn JN, Tavazzi L. Effect of valsartan on quality of life when added to usual therapy for heart failure: results from the Valsartan Heart Failure Trial. J. Card. Fail. 2005;11:253–259.

59. Edes I, Gasior Z, Wita K. Effects of nebivolol on left ventricular function in elderly patients with chronic heart failure: results of the ENECA study. Eur. J. Heart Fail. 2005;7:631–639.

60. Chan AKY, Sanderson JE, Wang T, Lam W, Yip G, Wang M, Lam Y-Y, Zhang Y, Yeung L, Wu EB, et al. Aldosterone receptor antagonism induces reverse remodeling when added to angiotensin receptor blockade in chronic heart failure. J. Am. Coll. Cardiol. 2007;50:591–596.

61. Ekman I, Chassany O, Komajda M, Böhm M, Borer JS, Ford I, Tavazzi L, Swedberg K. Heart rate reduction with ivabradine and health related quality of life in patients with chronic heart failure: results from the SHIFT study. Eur. Heart J. 2011;32:2395–2404.

62. Abdel-Salam Z, Rayan M, Saleh A, Abdel-Barr MG, Hussain M, Nammas W. I(f) current inhibitor ivabradine in patients with idiopathic dilated cardiomyopathy: Impact on the exercise tolerance and quality of life. Cardiol. J. 2015;22:227–232.

63. Lewis EF, Claggett BL, McMurray JJV, Packer M, Lefkowitz MP, Rouleau JL, Liu J, Shi VC, Zile MR, Desai AS, et al. Health-Related Quality of Life Outcomes in PARADIGM-HF. Circ. Heart Fail. 2017;10:e003430.

64. McMurray John J.V., Solomon Scott D., Inzucchi Silvio E., Køber Lars, Kosiborod Mikhail N., Martinez Felipe A., Ponikowski Piotr, Sabatine Marc S., Anand Inder S., Bělohlávek Jan, et al. Dapagliflozin in Patients with Heart Failure and Reduced Ejection Fraction. N. Engl. J. Med. 2019;381:1995–2008.

65. Felker GM, Solomon SD, McMurray JJV, Cleland JGF, Abbasi SA, Malik FI, Zhang H, Globe, Gary, Teerlink JR, Null N. Effects of Omecamtiv Mecarbil on Symptoms and Health-Related Quality of Life in Patients With Chronic Heart Failure. Circ. Heart Fail. 2020;13:e007814.

66. Teerlink John R., Diaz Rafael, Felker G. Michael, McMurray John J.V., Metra Marco, Solomon Scott D., Adams Kirkwood F., Anand Inder, Arias-Mendoza Alexandra, Biering-Sørensen Tor, et al. Cardiac Myosin Activation with Omecamtiv Mecarbil in Systolic Heart Failure. N. Engl. J. Med. 2021;384:105–116.

67. Tsutsui H, Momomura S-I, Saito Y, Ito H, Yamamoto K, Sakata Y, Desai AS, Ohishi T, Iimori T, Kitamura T, et al. Efficacy and Safety of Sacubitril/Valsartan in Japanese Patients With Chronic Heart Failure and Reduced Ejection Fraction - Results From the PARALLEL-HF Study. Circ. J. 2021;85:584–594.

68. Khandwalla RM, Grant D, Birkeland K, Heywood JT, Fombu E, Owens RL, Steinhubl SR, AWAKE-H. F. Study Investigators. The AWAKE-HF Study: Sacubitril/Valsartan Impact on Daily Physical Activity and Sleep in Heart Failure. Am. J. Cardiovasc. Drugs. 2021;21:241–254.

69. Halle M, Schöbel C, Winzer EB, Bernhardt P, Mueller S, Sieder C, Lecker LSM. A randomized clinical trial on the short-term effects of 12-week sacubitril/valsartan vs. enalapril on peak oxygen consumption in patients with heart failure with reduced ejection fraction: results from the ACTIVITY-HF study. Eur. J. Heart Fail. 2021;23:2073–2082.

70. Mann DL, Givertz MM, Vader JM, Starling RC, Shah P, McNulty SE, Anstrom KJ, Margulies KB, Kiernan MS, Mahr C, et al. Effect of Treatment With Sacubitril/Valsartan in Patients With Advanced Heart Failure and Reduced Ejection Fraction: A Randomized Clinical Trial. JAMA Cardiol. 2022;7:17–25.

71. Ye F, Wang X, Wu S, Ma S, Zhang Y, Liu G, Liu K, Yang Z, Pang X, Xue L, et al. Sustained-Release Ivabradine Hemisulfate in Patients With Systolic Heart Failure. J. Am. Coll. Cardiol. 2022;80:584–594.

72. Lee MMY, Brooksbank KJM, Wetherall K, Mangion K, Roditi G, Campbell RT, Berry C, Chong V, Coyle L, Docherty KF, et al. Effect of Empagliflozin on Left Ventricular Volumes in Patients With Type 2 Diabetes, or Prediabetes, and Heart Failure With Reduced Ejection Fraction (SUGAR-DM-HF). Circulation. 2021;143:516–525.

73. Spertus JA, Jones PG, Sandhu AT, Arnold SV. Interpreting the Kansas City Cardiomyopathy Questionnaire in Clinical Trials and Clinical Care: JACC State-of-the-Art Review. J. Am. Coll. Cardiol. 2020;76:2379–2390.

74. McDonald M, Virani S, Chan M, Ducharme A, Ezekowitz JA, Giannetti N, Heckman GA, Howlett JG, Koshman SL, Lepage S, et al. CCS/CHFS Heart Failure Guidelines Update: Defining a New Pharmacologic Standard of Care for Heart Failure With Reduced Ejection Fraction. Can. J. Cardiol. 2021;37:531–546.

75. Suebsaicharoen T, Chunekamrai P, Yingchoncharoen T, Tansawet A, Issarawattana T, Numthavaj P, Thakkinstian A. Comparative cardiovascular outcomes of novel drugs as an addition to conventional triple therapy for heart failure with reduced ejection fraction (HFrEF): a network meta-analysis of randomised controlled trials. Open Heart. 2023;10:e002364.

76. Xiang B, Yu Z, Zhou X. Comparative Efficacy of Medical Treatments for Chronic Heart Failure: A Network Meta-Analysis. Front Cardiovasc Med. 2021;8:787810.

77. De Marzo V, Savarese G, Tricarico L, Hassan S, Iacoviello M, Porto I, Ameri P. Network meta-analysis of medical therapy efficacy in more than 90,000 patients with heart failure and reduced ejection fraction. J. Intern. Med. 2022;292:333–349.

78. Fielding S, Ogbuagu A, Sivasubramaniam S, MacLennan G, Ramsay CR. Reporting and dealing with missing quality of life data in RCTs: has the picture changed in the last decade? Qual. Life Res. 2016;25:2977–2983.

79. Biering K, Hjollund NH, Frydenberg M. Using multiple imputation to deal with missing data and attrition in longitudinal studies with repeated measures of patient-reported outcomes. Clin. Epidemiol. 2015;7:91–106.

80. Digitalis Investigation Group. The Effect of Digoxin on Mortality and Morbidity in Patients with Heart Failure. N. Engl. J. Med. 1997;336:525–533.

81. Patocka J, Nepovimova E, Wu W, Kuca K. Digoxin: Pharmacology and toxicology-A review. Environ. Toxicol. Pharmacol. 2020;79:103400.

