## Supplementary Material for "A Network Meta-Analysis of Quality of Life in Heart Failure with Reduced Ejection Fraction"

### Supplemental Appendix

#### Supplemental Tables:

**Supplemental Table 1**. Risk of bias 2 assessment domain and overall scores for all included studies.

| **Unique ID** | **Experimental** | **Comparator** | **Study Design** | **D1** | **D2** | **D3** | **D4** | **D5** | **Overall** |
|---|---|---|---|---|---|---|---|---|---|
| Pollock et al. 1990 | BB | Placebo | DB, SC | 0 | 0 | 1 | 0 | 0 | Some Concerns |
| Widimsky et al. 1995 | ACEi | Placebo | DB, MC | 0 | 0 | 0 | 0 | 0 | Low risk |
| Bristow et al. 1996 | BB | Placebo | DB, MC | 0 | 0 | 0 | 0 | 0 | Low risk |
| Packer et al. 1996 | BB | Placebo | DB, MC | 0 | 0 | 0 | 0 | 0 | Low risk |
| Colucci et al. 1996 | BB | Placebo | DB, MC | 0 | 0 | 0 | 0 | 0 | Low risk |
| Cohn et al. 1997 | BB | Placebo | DB, MC | 0 | 0 | 0 | 0 | 0 | Low risk |
| Goldstein et al. 1999 | BB | Placebo | DB, MC | 0 | 0 | 1 | 0 | 0 | Some Concerns |
| Hjalmarson et al. 2000 | BB | Placebo | DB, MC | 0 | 0 | 1 | 0 | 0 | Some Concerns |
| Granger et al. 2000 | ARB | Placebo | DB, MC | 0 | 0 | 1 | 0 | 0 | Some Concerns |
| Beanlands et al. 2000 | BB | Placebo | DB, MC | 0 | 0 | 0 | 0 | 0 | Low Risk |
| De Milliano | BB | Placebo | DB, MC | 1 | 0 | 0 | 0 | 0 | Some Concerns |

| | | | | | | | | | |
|---|---|---|---|---|---|---|---|---|---|
| et al. 2001 | | | | | | | | | |
| Hutcheon et al. 2002 | ACEi | Placebo | DB, SC | 0 | 0 | 0 | 0 | 0 | Low Risk |
| Willenheimer et al. 2002 | ACEi | ARB | DB, MC | 0 | 0 | 2 | 0 | 0 | Major Concerns |
| Lader et al. 2003 | DIG | Placebo | DB, MC | 0 | 0 | 0 | 0 | 0 | Low Risk |
| Taylor et al. 2004 | H-ISDN | Placebo | DB, MC | 0 | 0 | 0 | 0 | 0 | Low Risk |
| Majani et al. 2004 | ARB | Placebo | DB, MC | 0 | 0 | 0 | 0 | 0 | Low Risk |
| Edes et al. 2005 | BB | Placebo | DB, MC | 0 | 0 | 0 | 0 | 0 | Low Risk |
| Chan et al. 2007 | ARB+MRA | ARB | DB, SC | 0 | 0 | 0 | 0 | 0 | Low Risk |
| Ekman et al. 2011 | Ivabradine | Placebo | DB, MC | 0 | 0 | 0 | 0 | 0 | Low Risk |
| Abdel-Salam et al. 2015 | Ivabradine | Placebo | DB, SC | 0 | 0 | 0 | 0 | 0 | Low Risk |
| Lewis et al. 2017 | ARNi | ACEi | DB, MC | 0 | 0 | 0 | 0 | 0 | Low Risk |
| Nassif et al. 2019 | SGLT2i | Placebo | DB, MC | 0 | 0 | 0 | 0 | 0 | Low Risk |
| Desai et al. 2019 | ARNi | ACEi | DB, MC | 0 | 0 | 0 | 0 | 0 | Low Risk |
| McMurray et al. 2019 | SGLT2i | Placebo | DB, MC | 0 | 0 | 0 | 0 | 0 | Low Risk |
| Jensen et al. 2020 | SGLT2i | Placebo | DB, MC | 0 | 0 | 0 | 0 | 0 | Low Risk |
| Felker et al. 2020 | OM | Placebo | DB, MC | 0 | 0 | 0 | 0 | 0 | Low Risk |
| Abraham et al. 2021 | SGLT2i | Placebo | DB, MC | 0 | 0 | 0 | 0 | 0 | Low Risk |

| | | | | | | | | | |
|---|---|---|---|---|---|---|---|---|---|
| Santos-Gallego et al. 2021 | SGLT2i | Placebo | DB, SC | 0 | 0 | 0 | 0 | 0 | Low Risk |
| Tsutsui et al. 2021 | ARNi | ACEi | DB, MC | 0 | 0 | 0 | 0 | 0 | Low Risk |
| Khandwalla et al. 2020 | ARNi | ACEi | DB, MC | 0 | 0 | 1 | 0 | 0 | Some Concerns |
| Lee et al. 2021 | SGLT2i | Placebo | DB, MC | 1 | 0 | 2 | 0 | 0 | Major Concerns |
| Teerlink et al. 2021 | OM | Placebo | DB, MC | 0 | 0 | 0 | 0 | 0 | Low Risk |
| Butler et al. 2021 | SGLT2i | Placebo | DB, MC | 0 | 0 | 1 | 0 | 0 | Some Concerns |
| Halle et al 2021 | ARNi | ACEi | DB, MC | 0 | 0 | 0 | 0 | 0 | Low Risk |
| Mann et al.. 2021 | ARNi | ARB | DB, MC | 0 | 0 | 1 | 0 | 0 | Some Concerns |
| Ye et al. 2022 | Ivabradine | Placebo | DB, MC | 0 | 0 | 0 | 0 | 0 | Low Risk |
| Palau et al. 2022 | SGLT2i | Placebo | DB, MC | 0 | 0 | 1 | 0 | 0 | Some Concerns |
| Spertus et al. 2022 | SGLT2i | Placebo | DB, MC | 0 | 0 | 0 | 0 | 0 | Low Risk |
| Butler et al. 2022 | Vericiguat | Placebo | DB, MC | 0 | 0 | 0 | 0 | 0 | Low Risk |
| Lewis et al. 2022 | OM | Placebo | DB, MC | 0 | 0 | 0 | 0 | 0 | Low Risk |
| McMurray et al | SGLT2i | Placebo | DB, MC | 0 | 0 | 0 | 0 | 0 | Low Risk |

Abbreviations: ACEI, angiotensin converting enzyme inhibitor; ARB, angiotensin-receptor blocker; ARNI, angiotensin receptor neprilysin inhibitor; BB, beta-blocker; DIG, digoxin; H-ISDN, hydralazine–isosorbide dinitrate; MRA, mineralocorticoid receptor antagonist; DB, double blind; MC, multi center; SC, single center.

**Supplemental Table 2**. Estimates of relative differences in treatment effect on quality of life of treatment combinations. The estimates presented are for the treatment combination in the column versus the treatment combination in the respective row.

| | **ACEi + BB + DIG + H-ISDN** | **ACEi + BB + MRA** | **ACEi + BB + MRA + Ivabradine** | **ACEi + BB + MRA + SGLT2i** | **ACEi + BB + MRA + Vericiguat** | **ARNi + BB + MRA** | **ARNi + BB + MRA + SGLT2i** | **ARNi + BB + SGLT2i** |
|---|---|---|---|---|---|---|---|---|
| **ACEi** | -1.23 (-8.32-5.86); 0.7345 | 2.03 (-4.75-8.82); 0.5575 | 5.26 (-2.17-12.69); 0.1653 | 5.34 (-1.69-12.36); 0.1364 | 3.03 (-4.81-10.87); 0.4487 | 3.83 (-3.34-11); 0.2957 | 7.13 (-0.26-14.52); 0.0587 | 5.34 (1.7-8.99); 0.0041 |
| **ACEi + ARB + DIG** | 4.11 (-1.92-10.13); 0.1818 | 7.36 (-1.42-16.14); 0.1002 | 10.59 (1.3-19.88); 0.0254 | 10.67 (1.7-19.63); 0.0197 | 8.36 (-1.26-17.98); 0.0884 | 9.16 (0.13-18.18); 0.0467 | 12.46 (3.26-21.67); 0.008 | 10.67 (4.09-17.26); 0.0015 |
| **ACEi + BB** | -1.47 (-8.22-5.29); 0.67 | 1.79 (-4.64-8.22); 0.5857 | 5.02 (-2.09-12.13); 0.1666 | 5.09 (-1.59-11.78); 0.1351 | 2.79 (-4.75-10.33); 0.4683 | 3.58 (-3.25-10.42); 0.3043 | 6.89 (-0.18-13.96); 0.0562 | 5.1 (2.16-8.04); 7e-04 |
| **ACEi + BB + DIG** | 3.87 (-0.94-8.68); 0.1146 | 7.13 (-0.86-15.12); 0.0804 | 10.36 (1.81-18.91); 0.0175 | 10.43 (2.24-18.63); 0.0126 | 8.13 (-0.78-17.04); 0.0736 | 8.92 (0.6-17.25); 0.0356 | 12.23 (3.71-20.75); 0.0049 | 10.44 (4.86-16.02); 2e-04 |
| **ACEi + BB + DIG + H-ISDN** | 1 | 3.26 (-6.07-12.59); 0.4937 | 6.49 (-3.32-16.3); 0.1948 | 6.56 (-2.94-16.06); 0.1758 | 4.26 (-5.86-14.38); 0.4097 | 5.05 (-4.56-14.67); 0.3029 | 8.36 (-1.42-18.14); 0.094 | 6.57 (-0.8-13.93); 0.0805 |
| **ACEi + BB + MRA** | -3.26 (-12.59-6.07); 0.4937 | 1 | 3.23 (0.2-6.26); 0.0368 | 3.3 (1.5-5.11); 3e-04 | 1 (-2.93-4.93); 0.6179 | 1.8 (-0.52-4.11); 0.1292 | 5.1 (2.16-8.04); 7e-04 | 3.31 (-3.76-10.38); 0.3589 |
| **ACEi + BB + MRA + Ivabradi** | -6.49 (-16.3-3.32); 0.1948 | -3.23 (-6.26--0.2); 0.0368 | 1 | 0.07 (-3.45-3.6); 0.9672 | -2.23 (-7.19-2.73); 0.3783 | -1.44 (-5.25-2.38); 0.4612 | 1.87 (-2.35-6.09); 0.3855 | 0.08 (-7.61-7.78); 0.9837 |

| | | | | | | | | |
|---|---|---|---|---|---|---|---|---|
| **ne** | | | | | | | | |
| **ACEi + BB + MRA + SGLT2i** | -6.56 (-16.06-2.94); 0.1758 | -3.3 (-5.11--1.5); 3e-04 | -0.07 (-3.6-3.45); 0.9672 | 1 | -2.3 (-6.63-2.02); 0.2961 | -1.51 (-4.45-1.43); 0.314 | 1.8 (-0.52-4.11); 0.1292 | 0.01 (-6.83-6.84); 0.9986 |
| **ACEi + BB + MRA + Vericiguat** | -4.26 (-14.38-5.86); 0.4097 | -1 (-4.93-2.93); 0.6179 | 2.23 (-2.73-7.19); 0.3783 | 2.3 (-2.02-6.63); 0.2961 | 1 | 0.8 (-3.77-5.36); 0.7326 | 4.1 (-0.81-9.01); 0.1014 | 2.31 (-5.78-10.4); 0.5756 |
| **ACEi + DIG** | 4.11 (-1.16-9.38); 0.1261 | 7.37 (-0.91-15.65); 0.081 | 10.6 (1.78-19.42); 0.0184 | 10.68 (2.2-19.15); 0.0135 | 8.37 (-0.79-17.53); 0.0734 | 9.17 (0.57-17.76); 0.0367 | 12.47 (3.69-21.26); 0.0054 | 10.68 (4.7-16.66); 5e-04 |
| **ARB + BB** | -1.49 (-9.03-6.04); 0.6977 | 1.76 (-5.48-9.01); 0.6332 | 5 (-2.86-12.85); 0.2127 | 5.07 (-2.4-12.54); 0.1834 | 2.76 (-5.48-11.01); 0.511 | 3.56 (-3.8-10.92); 0.3428 | 6.86 (-0.71-14.44); 0.0756 | 5.08 (1.08-9.07); 0.0128 |
| **ARB + BB + MRA** | -3.28 (-13.19-6.63); 0.5162 | -0.02 (-3.36-3.31); 0.9886 | 3.21 (-1.3-7.72); 0.1635 | 3.28 (-0.51-7.07); 0.0902 | 0.98 (-4.18-6.13); 0.7107 | 1.77 (-1.79-5.34); 0.3302 | 5.08 (1.08-9.07); 0.0128 | 3.29 (-4.29-10.86); 0.395 |
| **ARB + DIG** | 4.09 (-2.15-10.33); 0.1989 | 7.35 (-1.58-16.27); 0.1067 | 10.58 (1.15-20); 0.0279 | 10.65 (1.54-19.76); 0.0219 | 8.35 (-1.41-18.1); 0.0935 | 9.14 (0.13-18.16); 0.0468 | 12.45 (3.25-21.64); 0.008 | 10.66 (4.09-17.22); 0.0015 |
| **ARNi + BB** | -3.26 (-10.41-3.88); 0.3704 | -0.01 (-6.84-6.83); 0.9986 | 3.22 (-4.26-10.7); 0.3982 | 3.3 (-3.77-10.37); 0.3607 | 0.99 (-6.89-8.88); 0.805 | 1.79 (-4.64-8.22); 0.5857 | 5.09 (-1.59-11.78); 0.1351 | 3.3 (1.5-5.11); 3e-04 |
| **ARNi + BB + MRA** | -5.05 (-14.67-4.56); 0.3029 | -1.8 (-4.11-0.52); 0.1292 | 1.44 (-2.38-5.25); 0.4612 | 1.51 (-1.43-4.45); 0.314 | -0.8 (-5.36-3.77); 0.7326 | 1 | 3.3 (1.5-5.11); 3e-04 | 1.52 (-5.17-8.2); 0.6566 |
| **ARNi + BB + MRA + SGLT2i** | -8.36 (-18.14-1.42); 0.094 | -5.1 (-8.04--2.16); 7e-04 | -1.87 (-6.09-2.35); 0.3855 | -1.8 (-4.11-0.52); 0.1292 | -4.1 (-9.01-0.81); 0.1014 | -3.3 (-5.11--1.5); 3e-04 | 1 | -1.79 (-8.22-4.64); 0.5857 |

| | | | | | | | | |
|---|---|---|---|---|---|---|---|---|
| **ARNi + BB + SGLT2i** | -6.57 (-13.93-0.8); 0.0805 | -3.31 (-10.38-3.76); 0.3589 | -0.08 (-7.78-7.61); 0.9837 | -0.01 (-6.84-6.83); 0.9986 | -2.31 (-10.4-5.78); 0.5756 | -1.52 (-8.2-5.17); 0.6566 | 1.79 (-4.64-8.22); 0.5857 | 1 |
| **BB** | -1.49 (-9.2-6.22); 0.7058 | 1.77 (-5.66-9.2); 0.6401 | 5 (-3.02-13.03); 0.2217 | 5.08 (-2.57-12.72); 0.1931 | 2.77 (-5.63-11.18); 0.5179 | 3.57 (-4.03-11.17); 0.3576 | 6.87 (-0.94-14.68); 0.0847 | 5.08 (0.65-9.52); 0.0246 |
| **BB + DIG** | 3.86 (-2.22-9.93); 0.2138 | 7.11 (-1.7-15.93); 0.1138 | 10.34 (1.02-19.67); 0.0297 | 10.42 (1.42-19.42); 0.0233 | 8.11 (-1.54-17.76); 0.0994 | 8.91 (-0.05-17.87); 0.0513 | 12.21 (3.07-21.35); 0.0088 | 10.42 (3.93-16.92); 0.0017 |
| **DIG** | 4.1 (-2.35-10.55); 0.2131 | 7.35 (-1.72-16.43); 0.1122 | 10.58 (1.02-20.15); 0.0301 | 10.66 (1.41-19.91); 0.024 | 8.35 (-1.53-18.24); 0.0978 | 9.15 (-0.07-18.37); 0.0517 | 12.45 (3.06-21.85); 0.0093 | 10.67 (3.82-17.51); 0.0022 |
| **Placebo** | -1.24 (-9.25-6.76); 0.7608 | 2.01 (-5.72-9.75); 0.6099 | 5.24 (-3.07-13.55); 0.2161 | 5.32 (-2.63-13.26); 0.1894 | 3.01 (-5.66-11.69); 0.496 | 3.81 (-4.09-11.71); 0.3447 | 7.11 (-0.99-15.22); 0.0854 | 5.33 (0.4-10.25); 0.0342 |
| **SGLT2i** | -4.55 (-12.76-3.66); 0.2774 | -1.29 (-9.23-6.65); 0.7502 | 1.94 (-6.56-10.44); 0.6548 | 2.01 (-5.72-9.75); 0.6099 | -0.29 (-9.15-8.57); 0.9488 | 0.5 (-7.6-8.61); 0.9029 | 3.81 (-4.09-11.71); 0.3447 | 2.02 (-2.57-6.61); 0.388 |

Abbreviations: ACEI, angiotensin converting enzyme inhibitor; ARB, angiotensin-receptor blocker; ARNI, angiotensin receptor neprilysin inhibitor; BB, beta-blocker; DIG, digoxin; H-ISDN, hydralazine–isosorbide dinitrate; MRA, mineralocorticoid receptor antagonist.

**Supplemental Table 3**. Cinema framework domain summary for all included comparisons.

| Comparison | Number of studies | Within-study bias | Reporting bias | Indirectness | Imprecision | Heterogeneity | Incoherence | Confidence rating |
|---|---|---|---|---|---|---|---|---|
| ACEi:ACEi+BB | 1 | Some concerns | Low risk | No concerns | Major concerns | No concerns | No concerns | Low |
| ACEi:ACEi+DIG | 1 | No concerns | Low risk | No concerns | No concerns | No concerns | No concerns | High |
| ACEi:Placebo | 1 | No concerns | Low risk | Some concerns | Major concerns | No concerns | No concerns | Low |
| ACEi+ARB+DIG:ACEi+DIG | 1 | No concerns | Low risk | No concerns | No concerns | No concerns | No concerns | High |
| ACEi+BB:ARB+BB | 1 | Major concerns | Low risk | No concerns | No concerns | No concerns | No concerns | Low |
| ACEi+BB:ARNi+BB | 1 | No concerns | Low risk | No concerns | No concerns | No concerns | No concerns | High |
| ACEi+BB+DIG:ACEi+BB+DIG+H-ISDN | 1 | No concerns | Low risk | Some concerns | No concerns | No concerns | No concerns | Moderate |
| ACEi+BB+DIG:ACEi+DIG | 7 | No concerns | Low risk | No concerns | No concerns | No concerns | No concerns | High |
| ACEi+BB+MRA:ACEi+BB+MRA+Ivabradine | 3 | No concerns | Low risk | No concerns | No concerns | No concerns | No concerns | High |
| ACEi+BB+MRA:ACEi+BB+MRA+SGLT2i | 4 | No concerns | Low risk | No concerns | No concerns | No concerns | No concerns | High |
| ACEi+BB+MRA:ACEi+B | 1 | No concerns | Low risk | No concerns | No concerns | No concerns | No concerns | High |

| | | | | | | | | |
|---|---|---|---|---|---|---|---|---|
| B+MRA+Vericiguat | | | | | | | | |
| ACEi+BB+MRA:ARNi+BB+MRA | 3 | No concerns | Low risk | No concerns | No concerns | No concerns | No concerns | High |
| ACEi+DIG:DIG | 1 | No concerns | Low risk | No concerns | Some concerns | Some concerns | No concerns | Moderate |
| ARB+BB:ARB+BB+MRA | 1 | No concerns | Low risk | No concerns | Some concerns | No concerns | No concerns | Moderate |
| ARB+BB+MRA:ARNi+BB+MRA | 1 | Some concerns | Low risk | No concerns | No concerns | Some concerns | No concerns | Moderate |
| ARB+DIG:DIG | 1 | Some concerns | Low risk | No concerns | No concerns | No concerns | No concerns | Moderate |
| ARNi+BB:ARNi+BB+SGLT2i | 1 | No concerns | Low risk | No concerns | No concerns | No concerns | No concerns | High |
| ARNi+BB+MRA:ARNi+BB+MRA+SGLT2i | 2 | No concerns | Low risk | No concerns | No concerns | No concerns | No concerns | High |
| BB:Placebo | 1 | No concerns | Low risk | No concerns | Major concerns | No concerns | No concerns | Low |
| BB+DIG:DIG | 1 | Some concerns | Low risk | No concerns | No concerns | No concerns | No concerns | Moderate |
| Placebo:SGLT2i | 1 | No concerns | Low risk | No concerns | No concerns | Some concerns | No concerns | Moderate |
| ACEi:ACEi+ARB+DIG | 0 | No concerns | Low risk | No concerns | Some concerns | No concerns | No concerns | Moderate |
| ACEi:ACEi+BB+DIG | 0 | No concerns | Low risk | No concerns | No concerns | No concerns | No concerns | High |
| ACEi:ACEi+BB+DIG+H-ISDN | 0 | No concerns | Low risk | No concerns | Some concerns | No concerns | No concerns | Moderate |

| | | | | | | | | |
|---|---|---|---|---|---|---|---|---|
| ACEi:ACEi+BB+MRA | 0 | Some concerns | Low risk | No concerns | Major concerns | No concerns | No concerns | Low |
| ACEi:ACEi+BB+MRA+Ivabradine | 0 | Some concerns | Low risk | No concerns | Major concerns | No concerns | No concerns | Low |
| ACEi:ACEi+BB+MRA+SGLT2i | 0 | Some concerns | Low risk | No concerns | Major concerns | No concerns | No concerns | Low |
| ACEi:ACEi+BB+MRA+Vericiguat | 0 | Some concerns | Low risk | No concerns | Major concerns | No concerns | No concerns | Low |
| ACEi:ARB+BB | 0 | Major concerns | Low risk | No concerns | Major concerns | No concerns | No concerns | Very low |
| ACEi:ARB+BB+MRA | 0 | Some concerns | Low risk | No concerns | Major concerns | No concerns | No concerns | Low |
| ACEi:ARB+DIG | 0 | No concerns | Low risk | No concerns | No concerns | No concerns | No concerns | High |
| ACEi:ARNi+BB | 0 | Some concerns | Low risk | No concerns | Some concerns | Some concerns | No concerns | Low |
| ACEi:ARNi+BB+MRA | 0 | Some concerns | Low risk | No concerns | Major concerns | No concerns | No concerns | Low |
| ACEi:ARNi+BB+MRA+SGLT2i | 0 | Some concerns | Low risk | No concerns | Some concerns | Some concerns | No concerns | Low |
| ACEi:ARNi+BB+SGLT2i | 0 | No concerns | Low risk | No concerns | No concerns | No concerns | No concerns | High |
| ACEi:BB | 0 | No concerns | Low risk | Some concerns | Major concerns | No concerns | No concerns | Moderate |
| ACEi:BB+DIG | 0 | No concerns | Low risk | No concerns | Some concerns | No concerns | No concerns | Moderate |
| ACEi:DIG | 0 | No concerns | Low risk | No concerns | Some concerns | No concerns | No concerns | Moderate |
| ACEi:SGLT2i | 0 | No concerns | Low risk | Some concerns | Some concerns | No concerns | No concerns | Moderate |

| | | | | | | | | |
|---|---|---|---|---|---|---|---|---|
| ACEi+ARB+DIG:ACEi+BB | 0 | No concerns | Low risk | No concerns | Major concerns | No concerns | No concerns | Low |
| ACEi+ARB+DIG:ACEi+BB+DIG | 0 | No concerns | Low risk | No concerns | No concerns | Some concerns | No concerns | Moderate |
| ACEi+ARB+DIG:ACEi+BB+DIG+H-ISDN | 0 | No concerns | Low risk | No concerns | No concerns | Some concerns | No concerns | Moderate |
| ACEi+ARB+DIG:ACEi+BB+MRA | 0 | Some concerns | Low risk | No concerns | Major concerns | No concerns | No concerns | Low |
| ACEi+ARB+DIG:ACEi+BB+MRA+Ivabradine | 0 | Some concerns | Low risk | No concerns | Some concerns | Some concerns | No concerns | Low |
| ACEi+ARB+DIG:ACEi+BB+MRA+SGLT2i | 0 | Some concerns | Low risk | No concerns | Some concerns | Some concerns | No concerns | Low |
| ACEi+ARB+DIG:ACEi+BB+MRA+Vericiguat | 0 | Some concerns | Low risk | No concerns | Major concerns | No concerns | No concerns | Low |
| ACEi+ARB+DIG:ARB+BB | 0 | Some concerns | Low risk | No concerns | Major concerns | No concerns | No concerns | Low |
| ACEi+ARB+DIG:ARB+BB+MRA | 0 | Some concerns | Low risk | No concerns | Major concerns | No concerns | No concerns | Low |
| ACEi+ARB+DIG:ARB+DIG | 0 | No concerns | Low risk | No concerns | No concerns | No concerns | No concerns | High |
| ACEi+ARB+DIG:ARNi+BB | 0 | No concerns | Low risk | No concerns | Some concerns | No concerns | No concerns | Moderate |

| | | | | | | | | |
|---|---|---|---|---|---|---|---|---|
| ACEi+ARB+DIG:ARNi+BB+MRA | 0 | Some concerns | Low risk | No concerns | Major concerns | No concerns | No concerns | Low |
| ACEi+ARB+DIG:ARNi+BB+MRA+SGLT2i | 0 | Some concerns | Low risk | No concerns | Some concerns | No concerns | No concerns | Moderate |
| ACEi+ARB+DIG:ARNi+BB+SGLT2i | 0 | No concerns | Low risk | No concerns | No concerns | No concerns | No concerns | High |
| ACEi+ARB+DIG:BB | 0 | No concerns | Low risk | No concerns | Major concerns | No concerns | No concerns | Low |
| ACEi+ARB+DIG:BB+DIG | 0 | No concerns | Low risk | No concerns | No concerns | Some concerns | No concerns | Moderate |
| ACEi+ARB+DIG:DIG | 0 | No concerns | Low risk | No concerns | Some concerns | No concerns | No concerns | Moderate |
| ACEi+ARB+DIG:Placebo | 0 | No concerns | Low risk | No concerns | Some concerns | Some concerns | No concerns | Moderate |
| ACEi+ARB+DIG:SGLT2i | 0 | No concerns | Low risk | No concerns | Some concerns | No concerns | No concerns | Moderate |
| ACEi+BB:ACEi+BB+DIG | 0 | No concerns | Low risk | No concerns | Some concerns | No concerns | No concerns | Moderate |
| ACEi+BB:ACEi+BB+DIG+H-ISDN | 0 | No concerns | Low risk | No concerns | Major concerns | No concerns | No concerns | Low |
| ACEi+BB:ACEi+BB+MRA | 0 | Some concerns | Low risk | No concerns | Major concerns | No concerns | No concerns | Low |
| ACEi+BB:ACEi+BB+MRA+Ivabradine | 0 | Some concerns | Low risk | No concerns | Some concerns | No concerns | No concerns | Moderate |
| ACEi+BB:ACEi+BB+MRA+SGLT2i | 0 | Some concerns | Low risk | No concerns | Some concerns | No concerns | No concerns | Moderate |

| | | | | | | | | |
|---|---|---|---|---|---|---|---|---|
| ACEi+BB:ACEi+BB+MRA+Vericiguat | 0 | Some concerns | Low risk | No concerns | Major concerns | No concerns | No concerns | Low |
| ACEi+BB:ACEi+DIG | 0 | Some concerns | Low risk | No concerns | Some concerns | No concerns | No concerns | Moderate |
| ACEi+BB:ARB+BB+MRA | 0 | Some concerns | Low risk | No concerns | Some concerns | Some concerns | No concerns | Low |
| ACEi+BB:ARB+DIG | 0 | Some concerns | Low risk | No concerns | No concerns | Some concerns | No concerns | Moderate |
| ACEi+BB:ARNi+BB+MRA | 0 | Some concerns | Low risk | No concerns | Some concerns | Some concerns | No concerns | Low |
| ACEi+BB:ARNi+BB+MRA+SGLT2i | 0 | Some concerns | Low risk | No concerns | Some concerns | No concerns | No concerns | Moderate |
| ACEi+BB:ARNi+BB+SGLT2i | 0 | No concerns | Low risk | No concerns | No concerns | No concerns | No concerns | High |
| ACEi+BB:BB | 0 | No concerns | Low risk | No concerns | Major concerns | No concerns | No concerns | Low |
| ACEi+BB:BB+DIG | 0 | Some concerns | Low risk | No concerns | Major concerns | No concerns | No concerns | Low |
| ACEi+BB:DIG | 0 | No concerns | Low risk | No concerns | Some concerns | No concerns | No concerns | Moderate |
| ACEi+BB:Placebo | 0 | Some concerns | Low risk | Some concerns | Major concerns | No concerns | No concerns | Very low |
| ACEi+BB:SGLT2i | 0 | No concerns | Low risk | No concerns | Major concerns | No concerns | No concerns | Low |
| ACEi+BB+DIG:ACEi+BB+MRA | 0 | Some concerns | Low risk | No concerns | Some concerns | Some concerns | No concerns | Low |
| ACEi+BB+DIG:ACEi+BB+MRA+Ivabradine | 0 | Some concerns | Low risk | No concerns | Some concerns | No concerns | No concerns | Moderate |

| | | | | | | | | |
|---|---|---|---|---|---|---|---|---|
| ACEi+BB+DIG:ACEi+BB+MRA+SGLT2i | 0 | Some concerns | Low risk | No concerns | Some concerns | No concerns | No concerns | Moderate |
| ACEi+BB+DIG:ACEi+BB+MRA+Vericiguat | 0 | Some concerns | Low risk | No concerns | Some concerns | No concerns | No concerns | Moderate |
| ACEi+BB+DIG:ARB+BB | 0 | Some concerns | Low risk | No concerns | Some concerns | Some concerns | No concerns | Low |
| ACEi+BB+DIG:ARB+BB+MRA | 0 | Some concerns | Low risk | No concerns | Some concerns | No concerns | No concerns | Moderate |
| ACEi+BB+DIG:ARB+DIG | 0 | No concerns | Low risk | No concerns | Some concerns | No concerns | No concerns | Moderate |
| ACEi+BB+DIG:ARNi+BB | 0 | No concerns | Low risk | No concerns | No concerns | Some concerns | No concerns | Moderate |
| ACEi+BB+DIG:ARNi+BB+MRA | 0 | Some concerns | Low risk | No concerns | Some concerns | No concerns | No concerns | Moderate |
| ACEi+BB+DIG:ARNi+BB+MRA+SGLT2i | 0 | Some concerns | Low risk | No concerns | No concerns | Some concerns | No concerns | Moderate |
| ACEi+BB+DIG:ARNi+BB+SGLT2i | 0 | No concerns | Low risk | No concerns | No concerns | No concerns | No concerns | High |
| ACEi+BB+DIG:BB | 0 | No concerns | Low risk | No concerns | Major concerns | No concerns | No concerns | Low |
| ACEi+BB+DIG:BB+DIG | 0 | No concerns | Low risk | No concerns | No concerns | No concerns | No concerns | High |
| ACEi+BB+DIG:DIG | 0 | No concerns | Low risk | No concerns | Major concerns | No concerns | No concerns | Low |
| ACEi+BB+DIG:Placebo | 0 | No concerns | Low risk | No concerns | Some concerns | No concerns | No concerns | Moderate |

| | | | | | | | | |
|---|---|---|---|---|---|---|---|---|
| ACEi+BB+DIG:SGLT2i | 0 | No concerns | Low risk | No concerns | No concerns | No concerns | No concerns | High |
| ACEi+BB+DIG+H-ISDN: ACEi+BB+MRA | 0 | Some concerns | Low risk | No concerns | Major concerns | No concerns | No concerns | Low |
| ACEi+BB+DIG+H-ISDN: ACEi+BB+MRA+Ivabradine | 0 | No concerns | Low risk | No concerns | Major concerns | No concerns | No concerns | Low |
| ACEi+BB+DIG+H-ISDN: ACEi+BB+MRA+SGLT2i | 0 | Some concerns | Low risk | No concerns | Major concerns | No concerns | No concerns | Low |
| ACEi+BB+DIG+H-ISDN: ACEi+BB+MRA+Vericiguat | 0 | No concerns | Low risk | No concerns | Major concerns | No concerns | No concerns | Low |
| ACEi+BB+DIG+H-ISDN: ACEi+DIG | 0 | No concerns | Low risk | Some concerns | No concerns | Some concerns | No concerns | Moderate |
| ACEi+BB+DIG+H-ISDN: ARB+BB | 0 | Some concerns | Low risk | No concerns | Major concerns | No concerns | No concerns | Low |
| ACEi+BB+DIG+H-ISDN: ARB+BB+MRA | 0 | Some concerns | Low risk | No concerns | Major concerns | No concerns | No concerns | Low |
| ACEi+BB+DIG+H-ISDN: ARB+DIG | 0 | No concerns | Low risk | No concerns | No concerns | No concerns | No concerns | High |
| ACEi+BB+DIG+H-ISDN: ARNi+BB | 0 | No concerns | Low risk | No concerns | Some concerns | No concerns | No concerns | Moderate |

| | | | | | | | | |
|---|---|---|---|---|---|---|---|---|
| ACEi+BB+DIG+H-ISDN: ARNi+BB+MRA | 0 | Some concerns | Low risk | No concerns | Major concerns | No concerns | No concerns | Low |
| ACEi+BB+DIG+H-ISDN: ARNi+BB+MRA+SGLT2i | 0 | Some concerns | Low risk | No concerns | Some concerns | Some concerns | No concerns | Low |
| ACEi+BB+DIG+H-ISDN: ARNi+BB+SGLT2i | 0 | No concerns | Low risk | No concerns | No concerns | No concerns | No concerns | High |
| ACEi+BB+DIG+H-ISDN: BB | 0 | No concerns | Low risk | No concerns | Major concerns | No concerns | No concerns | Low |
| ACEi+BB+DIG+H-ISDN: BB+DIG | 0 | No concerns | Low risk | No concerns | Some concerns | No concerns | No concerns | Moderate |
| ACEi+BB+DIG+H-ISDN: DIG | 0 | No concerns | Low risk | No concerns | Some concerns | No concerns | No concerns | Moderate |
| ACEi+BB+DIG+H-ISDN: Placebo | 0 | No concerns | Low risk | Some concerns | Major concerns | No concerns | No concerns | Low |
| ACEi+BB+DIG+H-ISDN: SGLT2i | 0 | No concerns | Low risk | No concerns | Some concerns | No concerns | No concerns | Moderate |
| ACEi+BB+MRA:ACEi+DIG | 0 | Some concerns | Low risk | No concerns | Some concerns | Some concerns | No concerns | Low |
| ACEi+BB+MRA:ARB+BB | 0 | No concerns | Low risk | No concerns | Some concerns | Some concerns | No concerns | Moderate |
| ACEi+BB+MRA:ARB+BB+MRA | 0 | Some concerns | Low risk | No concerns | No concerns | No concerns | No concerns | Moderate |

| | | | | | | | | |
|---|---|---|---|---|---|---|---|---|
| ACEi+BB+MRA:ARB+DIG | 0 | Some concerns | Low risk | No concerns | No concerns | Some concerns | No concerns | Moderate |
| ACEi+BB+MRA:ARNi+BB | 0 | Some concerns | Low risk | No concerns | Some concerns | Some concerns | No concerns | Low |
| ACEi+BB+MRA:ARNi+BB+MRA+SGLT2i | 0 | No concerns | Low risk | No concerns | No concerns | No concerns | No concerns | High |
| ACEi+BB+MRA:ARNi+BB+SGLT2i | 0 | Some concerns | Low risk | No concerns | No concerns | No concerns | No concerns | Moderate |
| ACEi+BB+MRA:BB | 0 | Some concerns | Low risk | No concerns | Major concerns | No concerns | No concerns | Low |
| ACEi+BB+MRA:BB+DIG | 0 | Some concerns | Low risk | No concerns | Major concerns | No concerns | No concerns | Low |
| ACEi+BB+MRA:DIG | 0 | Some concerns | Low risk | No concerns | Some concerns | Some concerns | No concerns | Low |
| ACEi+BB+MRA:Placebo | 0 | Some concerns | Low risk | No concerns | Major concerns | No concerns | No concerns | Low |
| ACEi+BB+MRA:SGLT2i | 0 | Some concerns | Low risk | No concerns | Major concerns | No concerns | No concerns | Low |
| ACEi+BB+MRA+Ivabradine:ACEi+BB+MRA+SGLT2i | 0 | No concerns | Low risk | No concerns | No concerns | No concerns | No concerns | High |
| ACEi+BB+MRA+Ivabradine:ACEi+BB+MRA+Vericiguat | 0 | No concerns | Low risk | No concerns | No concerns | No concerns | No concerns | High |
| ACEi+BB+MRA+Ivabradi | 0 | Some concerns | Low risk | No concerns | Some concerns | No concerns | No concerns | Moderate |

| | | | | | | | | |
|---|---|---|---|---|---|---|---|---|
| ne:ACEi+DIG | | | | | | | | |
| ACEi+BB+MRA+Ivabradine:ARB+BB | 0 | No concerns | Low risk | No concerns | Some concerns | No concerns | No concerns | Moderate |
| ACEi+BB+MRA+Ivabradine:ARB+BB+MRA | 0 | No concerns | Low risk | No concerns | Some concerns | No concerns | No concerns | Moderate |
| ACEi+BB+MRA+Ivabradine:ARB+DIG | 0 | Some concerns | Low risk | No concerns | No concerns | No concerns | No concerns | Moderate |
| ACEi+BB+MRA+Ivabradine:ARNi+BB | 0 | Some concerns | Low risk | No concerns | Major concerns | No concerns | No concerns | Low |
| ACEi+BB+MRA+Ivabradine:ARNi+BB+MRA | 0 | No concerns | Low risk | No concerns | No concerns | No concerns | No concerns | High |
| ACEi+BB+MRA+Ivabradine:ARNi+BB+MRA+SGLT2i | 0 | No concerns | Low risk | No concerns | Some concerns | No concerns | No concerns | Moderate |
| ACEi+BB+MRA+Ivabradine:ARNi+BB+SGLT2i | 0 | No concerns | Low risk | No concerns | No concerns | No concerns | No concerns | High |
| ACEi+BB+MRA+Ivabradine:BB | 0 | Some concerns | Low risk | No concerns | Major concerns | No concerns | No concerns | Low |
| ACEi+BB+MRA+Ivabradine:BB+DIG | 0 | Some concerns | Low risk | No concerns | Major concerns | No concerns | No concerns | Low |
| ACEi+BB+MRA+Ivabradine:DIG | 0 | Some concerns | Low risk | No concerns | Some concerns | No concerns | No concerns | Moderate |

| | | | | | | | | |
|---|---|---|---|---|---|---|---|---|
| ACEi+BB+MRA+Ivabradine:Placebo | 0 | Some concerns | Low risk | No concerns | Major concerns | No concerns | No concerns | Low |
| ACEi+BB+MRA+Ivabradine:SGLT2i | 0 | Some concerns | Low risk | No concerns | Major concerns | No concerns | No concerns | Low |
| ACEi+BB+MRA+SGLT2i: ACEi+BB+MRA+Vericiguat | 0 | No concerns | Low risk | No concerns | No concerns | No concerns | No concerns | High |
| ACEi+BB+MRA+SGLT2i: ACEi+DIG | 0 | Some concerns | Low risk | No concerns | Some concerns | No concerns | No concerns | Moderate |
| ACEi+BB+MRA+SGLT2i: ARB+BB | 0 | No concerns | Low risk | No concerns | Some concerns | No concerns | No concerns | Moderate |
| ACEi+BB+MRA+SGLT2i: ARB+BB+MRA | 0 | No concerns | Low risk | No concerns | Some concerns | No concerns | No concerns | Moderate |
| ACEi+BB+MRA+SGLT2i: ARB+DIG | 0 | Some concerns | Low risk | No concerns | No concerns | No concerns | No concerns | Moderate |
| ACEi+BB+MRA+SGLT2i: ARNi+BB | 0 | Some concerns | Low risk | No concerns | Major concerns | No concerns | No concerns | Low |
| ACEi+BB+MRA+SGLT2i: ARNi+BB+MRA | 0 | No concerns | Low risk | No concerns | No concerns | No concerns | No concerns | High |
| ACEi+BB+MRA+SGLT2i: ARNi+BB+MRA+SGLT2i | 0 | No concerns | Low risk | No concerns | Some concerns | No concerns | No concerns | Moderate |
| ACEi+BB+MRA+SGLT2i: | 0 | No concerns | Low risk | No concerns | No concerns | No concerns | No concerns | High |

| | | | | | | | | |
|---|---|---|---|---|---|---|---|---|
| ARNi+BB+SGLT2i | | | | | | | | |
| ACEi+BB+MRA+SGLT2i: BB | 0 | Some concerns | Low risk | No concerns | Major concerns | No concerns | No concerns | Low |
| ACEi+BB+MRA+SGLT2i: BB+DIG | 0 | Some concerns | Low risk | No concerns | Major concerns | No concerns | No concerns | Low |
| ACEi+BB+MRA+SGLT2i: DIG | 0 | Some concerns | Low risk | No concerns | Some concerns | No concerns | No concerns | Moderate |
| ACEi+BB+MRA+SGLT2i: Placebo | 0 | Some concerns | Low risk | No concerns | Major concerns | No concerns | No concerns | Low |
| ACEi+BB+MRA+SGLT2i: SGLT2i | 0 | Some concerns | Low risk | No concerns | Major concerns | No concerns | No concerns | Low |
| ACEi+BB+MRA+Vericiguat:ACEi+DIG | 0 | Some concerns | Low risk | No concerns | Some concerns | No concerns | No concerns | Moderate |
| ACEi+BB+MRA+Vericiguat:ARB+BB | 0 | No concerns | Low risk | No concerns | Some concerns | No concerns | No concerns | Moderate |
| ACEi+BB+MRA+Vericiguat:ARB+BB+MRA | 0 | No concerns | Low risk | No concerns | No concerns | Some concerns | No concerns | Moderate |
| ACEi+BB+MRA+Vericiguat:ARB+DIG | 0 | Some concerns | Low risk | No concerns | No concerns | No concerns | No concerns | Moderate |
| ACEi+BB+MRA+Vericiguat:ARNi+BB | 0 | Some concerns | Low risk | No concerns | Major concerns | No concerns | No concerns | Low |
| ACEi+BB+MRA+Vericigu | 0 | No concerns | Low risk | No concerns | No concerns | No concerns | No concerns | High |

| | | | | | | | | |
|---|---|---|---|---|---|---|---|---|
| at:ARNi+BB +MRA | | | | | | | | |
| ACEi+BB+MRA+Vericiguat:ARNi+BB +MRA+SGLT2i | 0 | No concerns | Low risk | No concerns | No concerns | No concerns | No concerns | High |
| ACEi+BB+MRA+Vericiguat:ARNi+BB +SGLT2i | 0 | No concerns | Low risk | No concerns | No concerns | No concerns | No concerns | High |
| ACEi+BB+MRA+Vericiguat:BB | 0 | Some concerns | Low risk | No concerns | Major concerns | No concerns | No concerns | Low |
| ACEi+BB+MRA+Vericiguat:BB+DIG | 0 | Some concerns | Low risk | No concerns | Major concerns | No concerns | No concerns | Low |
| ACEi+BB+MRA+Vericiguat:DIG | 0 | Some concerns | Low risk | No concerns | Some concerns | No concerns | No concerns | Moderate |
| ACEi+BB+MRA+Vericiguat:Placebo | 0 | Some concerns | Low risk | No concerns | Major concerns | No concerns | No concerns | Low |
| ACEi+BB+MRA+Vericiguat:SGLT2i | 0 | Some concerns | Low risk | No concerns | Major concerns | No concerns | No concerns | Low |
| ACEi+DIG:ARB+BB | 0 | Some concerns | Low risk | No concerns | Some concerns | Some concerns | No concerns | Low |
| ACEi+DIG:ARB+BB+MRA | 0 | Some concerns | Low risk | No concerns | Some concerns | No concerns | No concerns | Moderate |
| ACEi+DIG:ARB+DIG | 0 | Some concerns | Low risk | No concerns | Some concerns | No concerns | No concerns | Moderate |
| ACEi+DIG:ARNi+BB | 0 | No concerns | Low risk | No concerns | No concerns | Some concerns | No concerns | Moderate |

| | | | | | | | | |
|---|---|---|---|---|---|---|---|---|
| ACEi+DIG:ARNi+BB+MRA | 0 | Some concerns | Low risk | No concerns | Some concerns | No concerns | No concerns | Moderate |
| ACEi+DIG:ARNi+BB+MRA+SGLT2i | 0 | Some concerns | Low risk | No concerns | No concerns | Some concerns | No concerns | Moderate |
| ACEi+DIG:ARNi+BB+SGLT2i | 0 | No concerns | Low risk | No concerns | No concerns | No concerns | No concerns | High |
| ACEi+DIG:BB | 0 | No concerns | Low risk | No concerns | Major concerns | No concerns | No concerns | Low |
| ACEi+DIG:BB+DIG | 0 | Some concerns | Low risk | No concerns | No concerns | No concerns | No concerns | Moderate |
| ACEi+DIG:Placebo | 0 | No concerns | Low risk | Some concerns | Some concerns | No concerns | No concerns | Moderate |
| ACEi+DIG:SGLT2i | 0 | No concerns | Low risk | No concerns | No concerns | No concerns | No concerns | High |
| ARB+BB:ARB+DIG | 0 | Some concerns | Low risk | No concerns | No concerns | Some concerns | No concerns | Moderate |
| ARB+BB:ARNi+BB | 0 | Some concerns | Low risk | No concerns | No concerns | Some concerns | No concerns | Moderate |
| ARB+BB:ARNi+BB+MRA | 0 | Some concerns | Low risk | No concerns | Some concerns | No concerns | No concerns | Moderate |
| ARB+BB:ARNi+BB+MRA+SGLT2i | 0 | No concerns | Low risk | No concerns | Some concerns | No concerns | No concerns | Moderate |
| ARB+BB:ARNi+BB+SGLT2i | 0 | Some concerns | Low risk | No concerns | No concerns | No concerns | No concerns | Moderate |
| ARB+BB:BB | 0 | Some concerns | Low risk | No concerns | Major concerns | No concerns | No concerns | Low |
| ARB+BB:BB+DIG | 0 | Some concerns | Low risk | No concerns | Major concerns | No concerns | No concerns | Low |
| ARB+BB:DIG | 0 | Some concerns | Low risk | No concerns | Some concerns | Some concerns | No concerns | Low |

| | | | | | | | | |
|---|---|---|---|---|---|---|---|---|
| ARB+BB:Placebo | 0 | Some concerns | Low risk | No concerns | Major concerns | No concerns | No concerns | Low |
| ARB+BB:SGLT2i | 0 | Some concerns | Low risk | No concerns | Major concerns | No concerns | No concerns | Low |
| ARB+BB+MRA:ARB+DIG | 0 | Some concerns | Low risk | No concerns | No concerns | Some concerns | No concerns | Moderate |
| ARB+BB+MRA:ARNi+BB | 0 | Some concerns | Low risk | No concerns | Some concerns | No concerns | No concerns | Moderate |
| ARB+BB+MRA:ARNi+BB+MRA+SGLT2i | 0 | Some concerns | Low risk | No concerns | Some concerns | No concerns | No concerns | Moderate |
| ARB+BB+MRA:ARNi+BB+SGLT2i | 0 | Some concerns | Low risk | No concerns | No concerns | No concerns | No concerns | Moderate |
| ARB+BB+MRA:BB | 0 | Some concerns | Low risk | No concerns | Major concerns | No concerns | No concerns | Low |
| ARB+BB+MRA:BB+DIG | 0 | Some concerns | Low risk | No concerns | Major concerns | No concerns | No concerns | Low |
| ARB+BB+MRA:DIG | 0 | Some concerns | Low risk | No concerns | Some concerns | No concerns | No concerns | Moderate |
| ARB+BB+MRA:Placebo | 0 | Some concerns | Low risk | No concerns | Major concerns | No concerns | No concerns | Low |
| ARB+BB+MRA:SGLT2i | 0 | Some concerns | Low risk | No concerns | Major concerns | No concerns | No concerns | Low |
| ARB+DIG:ARNi+BB | 0 | No concerns | Low risk | No concerns | No concerns | No concerns | No concerns | High |
| ARB+DIG:ARNi+BB+MRA | 0 | Some concerns | Low risk | No concerns | No concerns | No concerns | No concerns | Moderate |
| ARB+DIG:ARNi+BB+MRA+SGLT2i | 0 | Some concerns | Low risk | No concerns | No concerns | No concerns | No concerns | Moderate |

| | | | | | | | | |
|---|---|---|---|---|---|---|---|---|
| ARB+DIG:ARNi+BB+SGLT2i | 0 | No concerns | Low risk | No concerns | No concerns | No concerns | No concerns | High |
| ARB+DIG:BB | 0 | No concerns | Low risk | No concerns | Major concerns | No concerns | No concerns | Low |
| ARB+DIG:BB+DIG | 0 | Some concerns | Low risk | No concerns | No concerns | No concerns | No concerns | Moderate |
| ARB+DIG:Placebo | 0 | No concerns | Low risk | No concerns | No concerns | No concerns | No concerns | High |
| ARB+DIG:SGLT2i | 0 | No concerns | Low risk | No concerns | No concerns | No concerns | No concerns | High |
| ARNi+BB:ARNi+BB+MRA | 0 | Some concerns | Low risk | No concerns | Major concerns | No concerns | No concerns | Low |
| ARNi+BB:ARNi+BB+MRA+SGLT2i | 0 | Some concerns | Low risk | No concerns | Major concerns | No concerns | No concerns | Low |
| ARNi+BB:BB | 0 | No concerns | Low risk | No concerns | Major concerns | No concerns | No concerns | Low |
| ARNi+BB:BB+DIG | 0 | No concerns | Low risk | No concerns | Major concerns | No concerns | No concerns | Low |
| ARNi+BB:DIG | 0 | No concerns | Low risk | No concerns | No concerns | Some concerns | No concerns | Moderate |
| ARNi+BB:Placebo | 0 | No concerns | Low risk | No concerns | Major concerns | No concerns | No concerns | Low |
| ARNi+BB:SGLT2i | 0 | No concerns | Low risk | No concerns | Major concerns | No concerns | No concerns | Low |
| ARNi+BB+MRA:ARNi+BB+SGLT2i | 0 | Some concerns | Low risk | No concerns | No concerns | No concerns | No concerns | Moderate |
| ARNi+BB+MRA:BB | 0 | Some concerns | Low risk | No concerns | Major concerns | No concerns | No concerns | Low |
| ARNi+BB+MRA:BB+DIG | 0 | Some concerns | Low risk | No concerns | Major concerns | No concerns | No concerns | Low |

| | | | | | | | | |
|---|---|---|---|---|---|---|---|---|
| ARNi+BB+MRA:DIG | 0 | Some concerns | Low risk | No concerns | Some concerns | No concerns | No concerns | Moderate |
| ARNi+BB+MRA:Placebo | 0 | Some concerns | Low risk | No concerns | Major concerns | No concerns | No concerns | Low |
| ARNi+BB+MRA:SGLT2i | 0 | Some concerns | Low risk | No concerns | Major concerns | No concerns | No concerns | Low |
| ARNi+BB+MRA+SGLT2i: ARNi+BB+SGLT2i | 0 | Some concerns | Low risk | No concerns | No concerns | No concerns | No concerns | Moderate |
| ARNi+BB+MRA+SGLT2i: BB | 0 | Some concerns | Low risk | No concerns | Major concerns | No concerns | No concerns | Low |
| ARNi+BB+MRA+SGLT2i: BB+DIG | 0 | Some concerns | Low risk | No concerns | Major concerns | No concerns | No concerns | Low |
| ARNi+BB+MRA+SGLT2i: DIG | 0 | Some concerns | Low risk | No concerns | No concerns | Some concerns | No concerns | Moderate |
| ARNi+BB+MRA+SGLT2i: Placebo | 0 | Some concerns | Low risk | No concerns | Major concerns | No concerns | No concerns | Low |
| ARNi+BB+MRA+SGLT2i: SGLT2i | 0 | Some concerns | Low risk | No concerns | Major concerns | No concerns | No concerns | Low |
| ARNi+BB+SGLT2i:BB | 0 | No concerns | Low risk | No concerns | No concerns | No concerns | No concerns | High |
| ARNi+BB+SGLT2i:BB+DIG | 0 | No concerns | Low risk | No concerns | Major concerns | No concerns | No concerns | Low |
| ARNi+BB+SGLT2i:DIG | 0 | No concerns | Low risk | No concerns | No concerns | No concerns | No concerns | High |
| ARNi+BB+SGLT2i:Placebo | 0 | No concerns | Low risk | No concerns | No concerns | No concerns | No concerns | High |

| | | | | | | | | |
|---|---|---|---|---|---|---|---|---|
| ARNi+BB+SGLT2i:SGLT2i | 0 | No concerns | Low risk | No concerns | No concerns | No concerns | No concerns | High |
| BB:BB+DIG | 0 | No concerns | Low risk | No concerns | Some concerns | Some concerns | No concerns | Moderate |
| BB:DIG | 0 | No concerns | Low risk | No concerns | Major concerns | No concerns | No concerns | Low |
| BB:SGLT2i | 0 | No concerns | Low risk | No concerns | Some concerns | No concerns | No concerns | Moderate |
| BB+DIG:Placebo | 0 | No concerns | Low risk | No concerns | Major concerns | No concerns | No concerns | Low |
| BB+DIG:SGLT2i | 0 | No concerns | Low risk | No concerns | Major concerns | No concerns | No concerns | Low |
| DIG:Placebo | 0 | No concerns | Low risk | No concerns | Some concerns | No concerns | No concerns | Moderate |
| DIG:SGLT2i | 0 | No concerns | Low risk | No concerns | No concerns | Some concerns | No concerns | Moderate |

Abbreviations: ACEI, angiotensin converting enzyme inhibitor; ARB, angiotensin-receptor blocker; ARNI, angiotensin receptor neprilysin inhibitor; BB, beta-blocker; DIG, digoxin; H-ISDN, hydralazine–isosorbide dinitrate; MRA, mineralocorticoid receptor antagonist.

**Supplemental Table 4**: *P*-scores of each component.

| **Treatment combination** | ***P*-score** |
|---|---:|
| ARNi + BB + MRA + SGLT2i | 0.948 |
| ARNi + BB + SGLT2i | 0.859 |
| ACEi + BB + MRA + SGLT2i | 0.842 |
| ACEi + BB + MRA + Ivabradine | 0.831 |
| SGLT2i | 0.739 |
| ARNi + BB + MRA | 0.727 |
| ACEi + BB + MRA + Vericiguat | 0.655 |
| ARNi + BB | 0.642 |
| ARB + BB + MRA | 0.579 |
| ACEi + BB + MRA | 0.566 |
| BB | 0.481 |
| ARB + BB | 0.48 |
| ACEi + BB | 0.473 |
| Placebo | 0.46 |
| ACEi | 0.454 |
| ACEi + BB + DIG + H-ISDN | 0.415 |
| BB + DIG | 0.154 |
| ACEi + BB + DIG | 0.149 |
| ACEi + ARB + DIG | 0.143 |
| ARB + DIG | 0.137 |
| DIG | 0.136 |
| ACEi + DIG | 0.131 |

Abbreviations: ACEI, angiotensin converting enzyme inhibitor; ARB, angiotensin-receptor blocker; ARNI, angiotensin receptor neprilysin inhibitor; BB, beta-blocker; DIG, digoxin; H-ISDN, hydralazine–isosorbide dinitrate; MRA, mineralocorticoid receptor antagonist.

**Supplemental Figures:**

**Supplemental Figure 1**. Funnel plots for quality of life (A) and discontinuation (B) outcomes

Abbreviations: ACEI, angiotensin converting enzyme inhibitor; ARB, angiotensin-receptor blocker; ARNI, angiotensin receptor neprilysin inhibitor; BB, beta-blocker; Dig, digoxin; H-ISDN, hydralazine–isosorbide dinitrate; MRA, mineralocorticoid receptor antagonist; SGLT2i, sodium-glucose cotransporter 2 inhibitor.; OM, omecamtiv mecarbil.

A

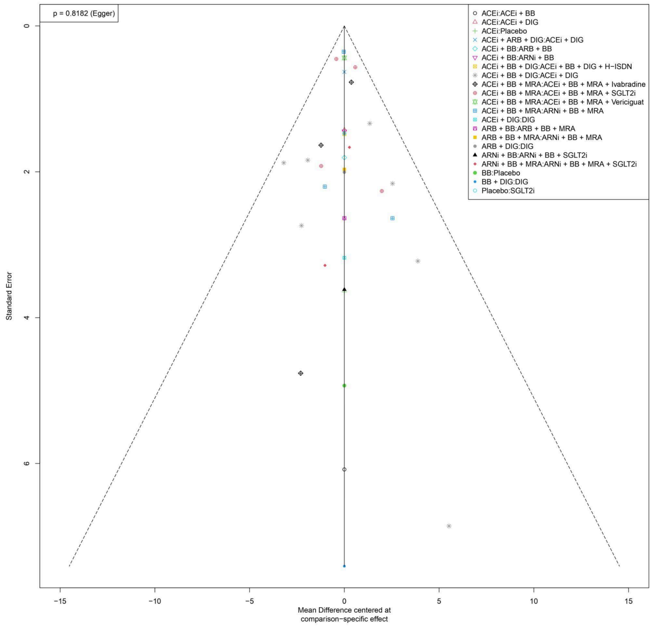


B

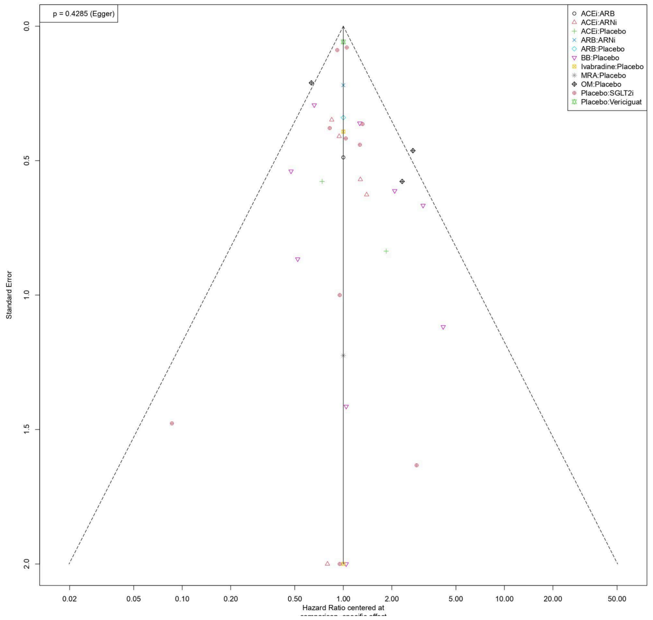

**Supplemental Figure 2**. Forest plot showing the risk of drug discontinuation of different treatment combinations against placebo.

Abbreviations: ACEI, angiotensin converting enzyme inhibitor; ARB, angiotensin-receptor blocker; ARNI, angiotensin receptor neprilysin inhibitor; BB, beta-blocker; DIG, digoxin; H-ISDN, hydralazine–isosorbide dinitrate; MRA, mineralocorticoid receptor antagonist; OM, omecamtiv mecarbil.

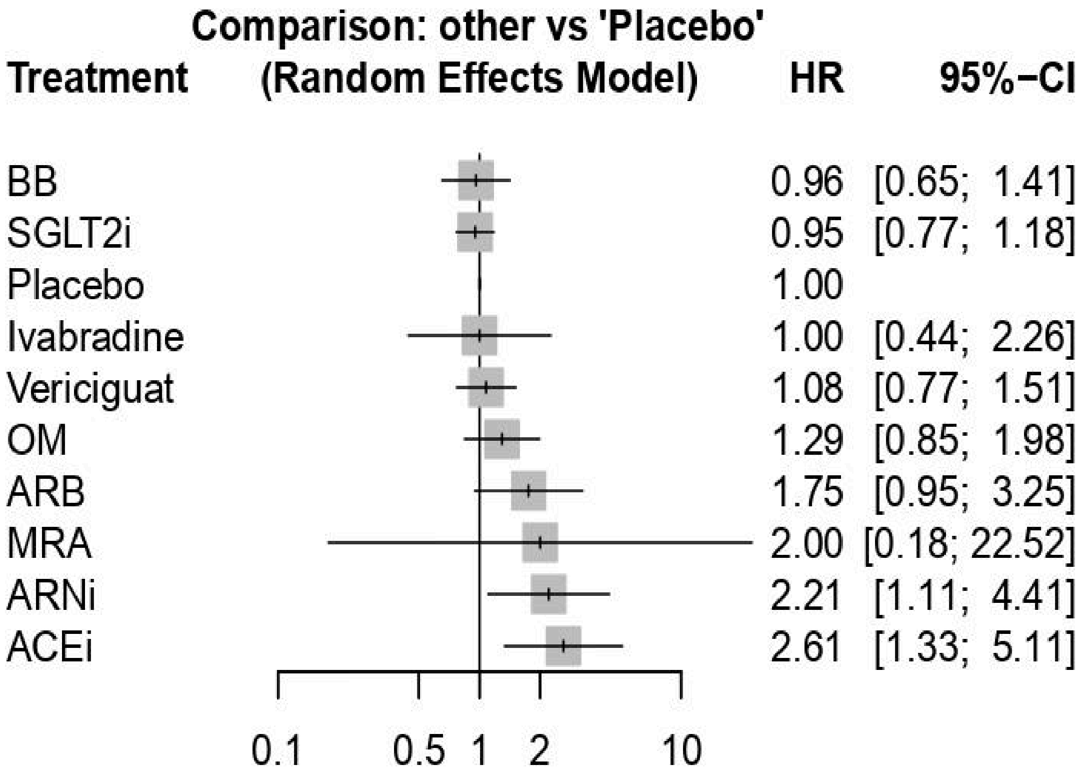

**Supplemental Figure 3**. Forest plot showing the mean difference in quality of life score change in various treatment combinations against placebo using a fixed effects model.

Abbreviations: ACEI, angiotensin converting enzyme inhibitor; ARB, angiotensin-receptor blocker; ARNI, angiotensin receptor neprilysin inhibitor; BB, beta-blocker; DIG, digoxin; H-ISDN, hydralazine–isosorbide dinitrate; MRA, mineralocorticoid receptor antagonist; OM, omecamtiv mecarbil.

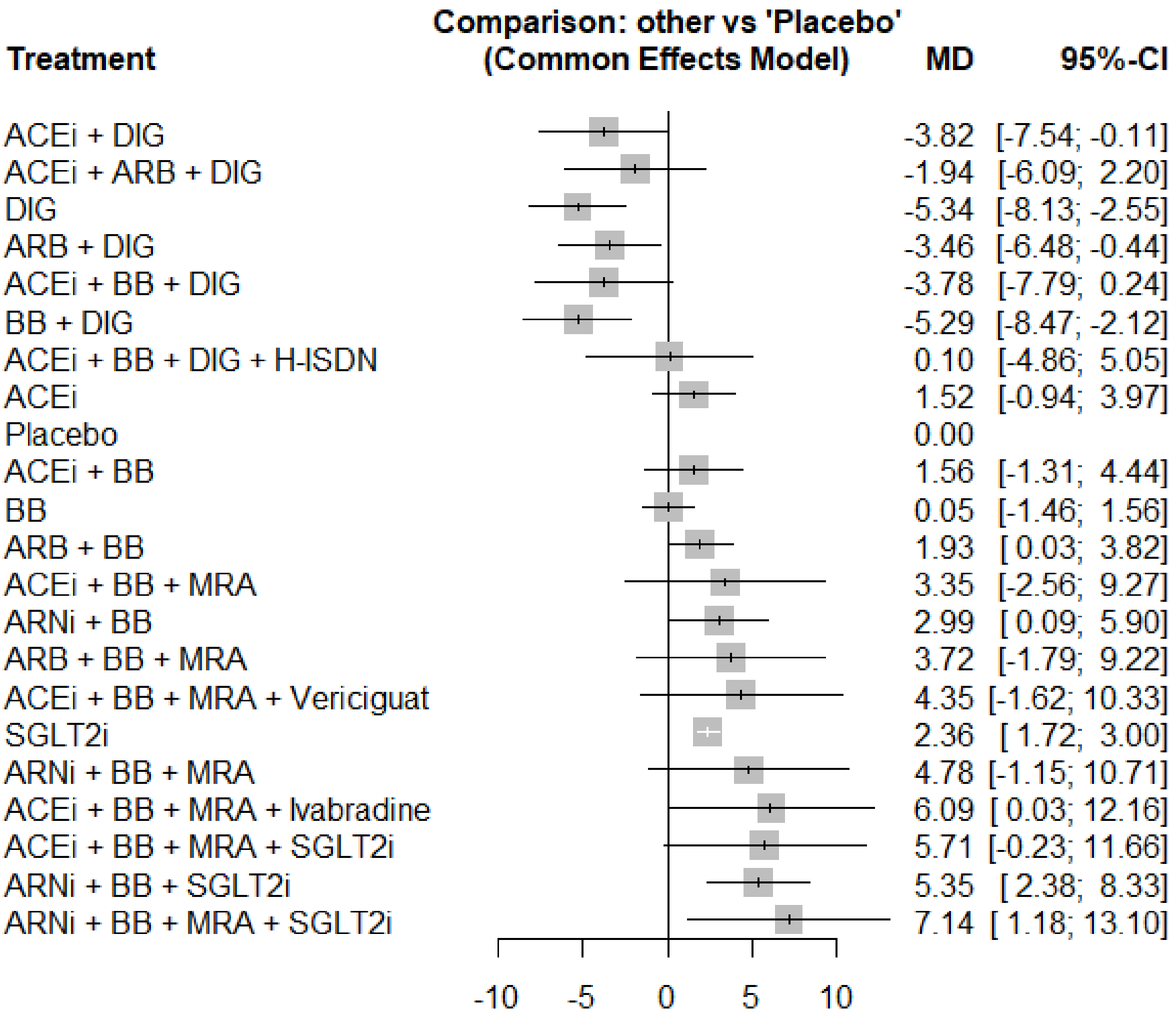

**Supplemental Figure 4.** Sensitivity analysis showing the mean difference in quality of life Score for various Treatments only in studies which used the MLHFQ score (A) and only in studies which used the MLHFQ score and were subsequently converted to KCCQ (B).

Abbreviations: ACEI, angiotensin converting enzyme inhibitor; ARB, angiotensin-receptor blocker; ARNI, angiotensin receptor neprilysin inhibitor; BB, beta-blocker; DIG, digoxin; H-ISDN, hydralazine–isosorbide dinitrate; MRA, mineralocorticoid receptor antagonist; SGLT2i, sodium-glucose cotransporter 2 inhibitor.

A

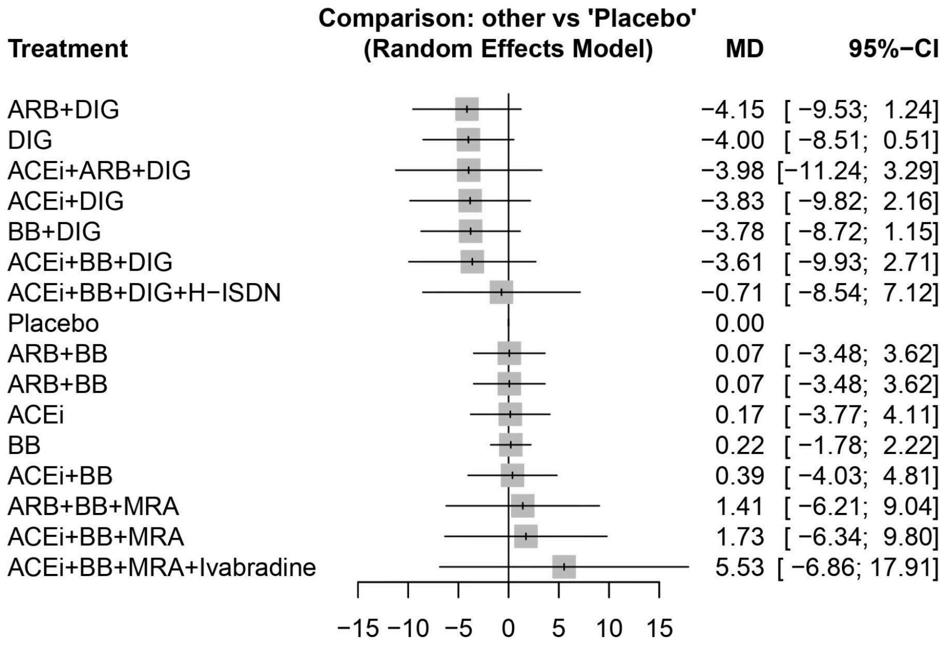


B

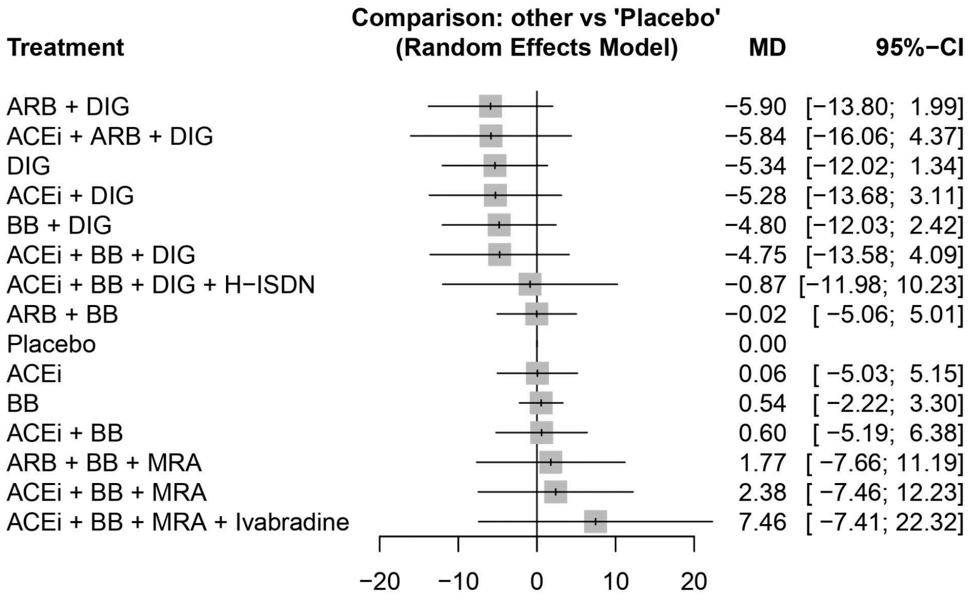

**Supplemental Figure 5**. Sensitivity analysis showing the mean difference in quality of life score for various treatments after the removal of studies with major concerns of bias.

Abbreviations: ACEI, angiotensin converting enzyme inhibitor; ARB, angiotensin-receptor blocker; ARNI, angiotensin receptor neprilysin inhibitor; BB, beta-blocker; DIG, digoxin; H-ISDN, hydralazine–isosorbide dinitrate; MRA, mineralocorticoid receptor antagonist.

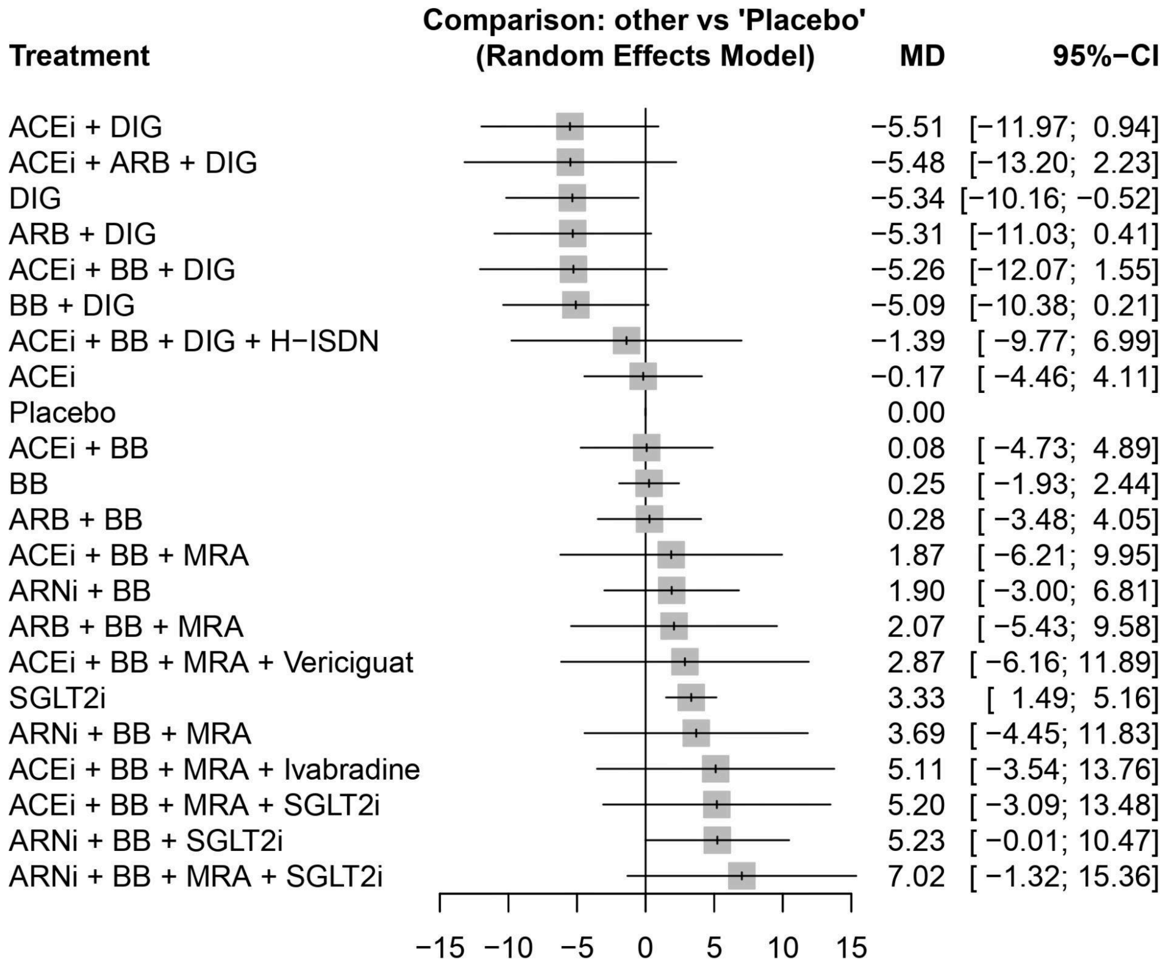

**Supplemental Figure 6**. Sensitivity analysis showing the mean difference in quality of life score change considering trials on a background of ACEi instead of ARNi.

Abbreviations: ACEI, angiotensin converting enzyme inhibitor; ARB, angiotensin-receptor blocker; ARNI, angiotensin receptor neprilysin inhibitor; BB, beta-blocker; DIG, digoxin; H-ISDN, hydralazine–isosorbide dinitrate; MRA, mineralocorticoid receptor antagonist; OM, omecamtiv mecarbil.

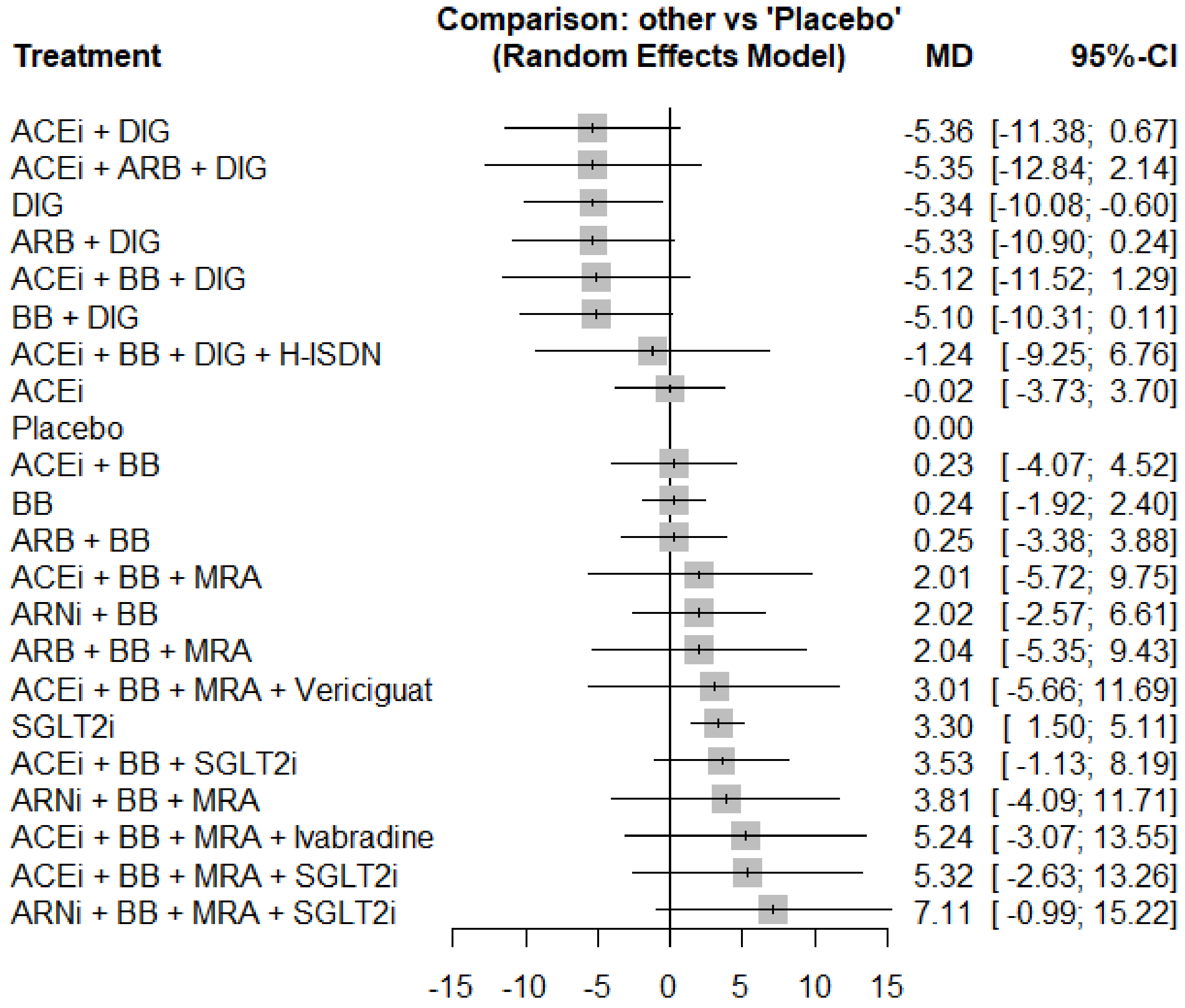

**Supplemental Figure 7**. Sensitivity analysis showing the mean difference in quality of life score change after the removal of studies which did not report background therapies.

Abbreviations: ACEI, angiotensin converting enzyme inhibitor; ARB, angiotensin-receptor blocker; ARNI, angiotensin receptor neprilysin inhibitor; BB, beta-blocker; DIG, digoxin; H-ISDN, hydralazine–isosorbide dinitrate; MRA, mineralocorticoid receptor antagonist; OM, omecamtiv mecarbil.

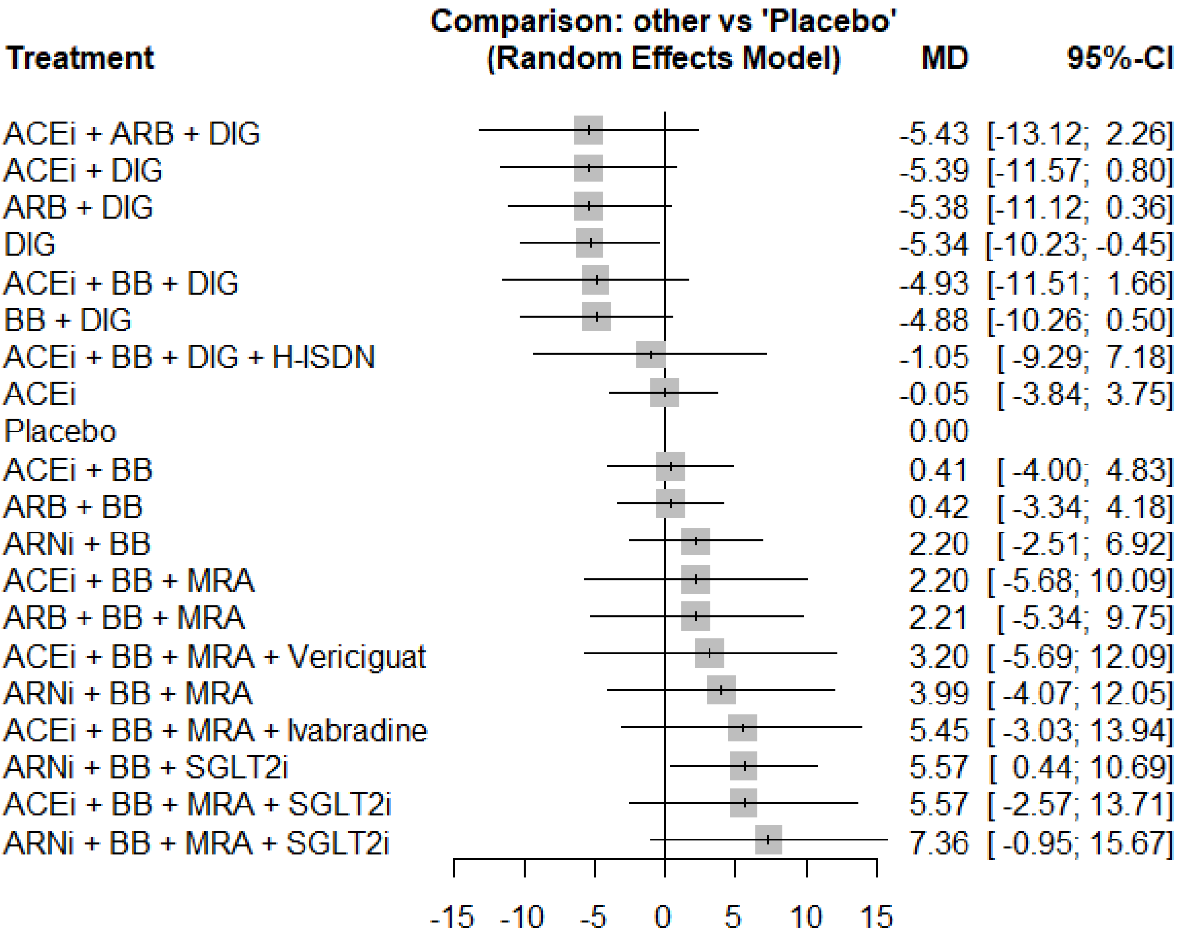

SEARCH TERMS:

***MEDLINE(R)/EMBASE search***
**Date of search:** Feb 28 2023
1. exp Heart Failure/
2. Cardiomyopathy, Dilated/
3. (heart failure or cardiac failure or cardiac insufficiency or cardiomyopath$).tw.
4. ((cardi$ or myocard$) adj2 (failure$ or insufficien$)).tw.
5. OR/1-4
6. exp angiotensin receptor-neprilysin inhibitor/ OR ARNI
7. LCZ696 OR LCZ 696 OR LCZ-696 OR valsartan adj6 sacubitril OR valsartan adj6 sacubitril OR valsartan sacubitril OR valsartan-sacubitril OR sacubitril adj6valsartan OR sacubitril valsartan OR sacubitril-valsartan
8. exp dipeptidyl carboxypeptidase inhibitor/ OR exp Angiotensin-Converting Enzyme Inhibitors/
9. (angiotensin converting enzyme inhibitor OR ACEI OR ACEI OR antagonist$ OR inhibitor$ benazepril OR captopril OR enalapril OR fosinopril OR imidapril OR lisinopril OR moexipril OR perindopril OR quinapril OR ramipril OR trandolapril OR zofenopril OR alacepril OR cilazapril OR spirapril OR delapril).mp.
10. exp beta adrenergic receptor blocking agent/ OR exp Adrenergic beta-Antagonists/
11. (beta blocker$ OR BB OR acebutolol OR atenolol OR betaxolol OR bisoprolol OR carvedilol OR labetalol OR metoprolol OR nadolol OR nebivolol OR penbutolol OR pindolol OR propranolol OR sotalol OR timolol).mp.
12. exp aldosterone antagonist/
13. (aldosterone antagonist$ OR mineralocorticoid-receptor antagonist OR MRA OR eplerenone OR spironolactone).mp.
14. exp angiotensin receptor antagonist/
15. (angiotensin receptor blocker$ OR angiotensin receptor antagonist$ OR ARB OR azilsartan OR candesartan OR eprosartan OR irbesartan OR losartan OR olmesartan OR telmisartan OR valsartan).mp.
16. sodium-glucose co-transporter 2 OR SGLT2 OR SGLT2 inhibitor* OR sodium glucose adj6 inhibitor* OR (SGLT2 inhibitor*) OR sodium-glucose adj6 inhibitor* OR Sodium-Glucose Transporter 2 OR sodium glucose-cotransporter 2 OR sodium-glucose co-transporter$ OR sodium glucose-cotransporter$
17. (dapagliflozin OR empagliflozin).mp.
18 exp ivabradine plus metoprolol/ or exp ivabradine/ or exp carvedilol plus ivabradine/
19 (Omecamtiv mecarbil OR CK-1827452 OR Omecamtiv OR mecarbil)
20 (Vericiguat OR guanylate cyclase stimulator OR guanylate cyclase stimulator)
21. (Hydralazine-Isosorbide Dinitrate OR Hydralazine-Isosorbide adj6 Dinitrate OR Hydralazine Isosorbide Dinitrate OR Hydralazine adj6 Isosorbide adj6 Dinitrate)
22. OR/6-21
23. "randomized controlled trial".pt.
24. (random$ or placebo$ or single blind$ or double blind$ or triple blind$).ti,ab.
25. (retraction of publication or retracted publication).pt.
26. OR/23-25
27. "Quality of Life"/
28. (kansas city cardiomyopathy questionnaire or kccq)
29. ("minnesota living with heart failure questionnaire" or mlhfq)

30. ("quality of life" or qol or hrqol)
31. OR/27-30
32. 22 AND 31
33. (animals not humans).sh.
34. ((comment or editorial or meta-analysis or practice-guideline or review or letter or journal correspondence) not "randomized controlled trial").pt.
35. (random sampl$ or random digit$ or random effect$ or random survey or random regression).ti,ab. not "randomized controlled trial".pt.
36. 33 OR 34 OR 35
37. 26 NOT 36
38. (random$ or placebo$ or single blind$ or double blind$ or triple blind$).ti,ab.
39. RETRACTED ARTICLE/
40. OR/38-39
41. (animal$ not human$).sh,hw.
42. (book or conference paper or editorial or letter or review).pt. not exp randomized controlled trial/
43. (random sampl$ or random digit$ or random effect$ or random survey or random regression).ti,ab. not exp randomized controlled trial/
44. OR/41-43
45. 40 NOT 44
46. 37 OR 45
47. 5 AND 32 AND 46
48. limit 47 to (human and yr="2020-Current" and (adult <18 to 64 years> or aged <65+ years>))

***MEDLINE(R)/EMBASE search***
**Date of search:** Feb 20 2024

1. exp Heart Failure/
2. Cardiomyopathy, Dilated/
3. (heart failure or cardiac failure or cardiac insufficiency or cardiomyopath$).tw.
4. ((cardi$ or myocard$) adj2 (failure$ or insufficien$)).tw.
5. OR/1-4
6. exp angiotensin receptor-neprilysin inhibitor/ OR ARNI
7. LCZ696 OR LCZ 696 OR LCZ-696 OR valsartan adj6 sacubitril OR valsartan adj6 sacubitril OR valsartan sacubitril OR valsartan-sacubitril OR sacubitril adj6valsartan OR sacubitril valsartan OR sacubitril-valsartan
8. exp dipeptidyl carboxypeptidase inhibitor/ OR exp Angiotensin-Converting Enzyme Inhibitors/
9. (angiotensin converting enzyme inhibitor OR ACEI OR ACEI OR antagonist$ OR inhibitor$ benazepril OR captopril OR enalapril OR fosinopril OR imidapril OR lisinopril OR moexipril OR perindopril OR quinapril OR ramipril OR trandolapril OR zofenopril OR alacepril OR cilazapril OR spirapril OR delapril).mp.
10. exp beta adrenergic receptor blocking agent/ OR exp Adrenergic beta-Antagonists/
11. (beta blocker$ OR BB OR acebutolol OR atenolol OR betaxolol OR bisoprolol OR carvedilol OR labetalol OR metoprolol OR nadolol OR nebivolol OR penbutolol OR pindolol OR propranolol OR sotalol OR timolol).mp.
12. exp aldosterone antagonist/

13. (aldosterone antagonist$ OR mineralocorticoid-receptor antagonist OR MRA OR eplerenone OR spironolactone).mp.
14. exp angiotensin receptor antagonist/
15. (angiotensin receptor blocker$ OR angiotensin receptor antagonist$ OR ARB OR azilsartan OR candesartan OR eprosartan OR irbesartan OR losartan OR olmesartan OR telmisartan OR valsartan).mp.
16. sodium-glucose co-transporter 2 OR SGLT2 OR SGLT2 inhibitor* OR sodium glucose adj6 inhibitor* OR (SGLT2 inhibitor*) OR sodium-glucose adj6 inhibitor* OR Sodium-Glucose Transporter 2 OR sodium glucose-cotransporter 2 OR sodium-glucose co-transporter$ OR sodium glucose-cotransporter$
17. (dapagliflozin OR empagliflozin).mp.
18 exp ivabradine plus metoprolol/ or exp ivabradine/ or exp carvedilol plus ivabradine/
19 (Omecamtiv mecarbil OR CK-1827452 OR Omecamtiv OR mecarbil)
20 (Vericiguat OR guanylate cyclase stimulator OR guanylate cyclase stimulator)
21. (Hydralazine-Isosorbide Dinitrate OR Hydralazine-Isosorbide adj6 Dinitrate OR Hydralazine Isosorbide Dinitrate OR Hydralazine adj6 Isosorbide adj6 Dinitrate)
22. OR/6-21
23. "randomized controlled trial".pt.
24. (random$ or placebo$ or single blind$ or double blind$ or triple blind$).ti,ab.
25. (retraction of publication or retracted publication).pt.
26. OR/23-25
27. "Quality of Life"/
28. (kansas city cardiomyopathy questionnaire or kccq)
29. ("minnesota living with heart failure questionnaire" or mlhfq)
30. ("quality of life" or qol or hrqol)
31. OR/27-30
32. 22 AND 31
33. (animals not humans).sh.
34. ((comment or editorial or meta-analysis or practice-guideline or review or letter or journal correspondence) not "randomized controlled trial").pt.
35. (random sampl$ or random digit$ or random effect$ or random survey or random regression).ti,ab. not "randomized controlled trial".pt.
36. 33 OR 34 OR 35
37. 26 NOT 36
38. (random$ or placebo$ or single blind$ or double blind$ or triple blind$).ti,ab.
39. RETRACTED ARTICLE/
40. OR/38-39
41. (animal$ not human$).sh,hw.
42. (book or conference paper or editorial or letter or review).pt. not exp randomized controlled trial/
43. (random sampl$ or random digit$ or random effect$ or random survey or random regression).ti,ab. not exp randomized controlled trial/
44. OR/41-43
45. 40 NOT 44
46. 37 OR 45
47. 5 AND 32 AND 46
48. limit 47 to (human and yr="2023-Current" and (adult <18 to 64 years> or aged <65+ years>))

***MEDLINE(R)/EMBASE search***
**Date of search:** Aug 10 2024

1. exp Heart Failure/
2. Cardiomyopathy, Dilated/
3. (heart failure or cardiac failure or cardiac insufficiency or cardiomyopath$).tw.
4. ((cardi$ or myocard$) adj2 (failure$ or insufficien$)).tw.
5. OR/1-4
6. exp angiotensin receptor-neprilysin inhibitor/ OR ARNI
7. LCZ696 OR LCZ 696 OR LCZ-696 OR valsartan adj6 sacubitril OR valsartan adj6 sacubitril OR valsartan sacubitril OR valsartan-sacubitril OR sacubitril adj6valsartan OR sacubitril valsartan OR sacubitril-valsartan
8. exp dipeptidyl carboxypeptidase inhibitor/ OR exp Angiotensin-Converting Enzyme Inhibitors/
9. (angiotensin converting enzyme inhibitor OR ACEI OR ACEI OR antagonist$ OR inhibitor$ benazepril OR captopril OR enalapril OR fosinopril OR imidapril OR lisinopril OR moexipril OR perindopril OR quinapril OR ramipril OR trandolapril OR zofenopril OR alacepril OR cilazapril OR spirapril OR delapril).mp.
10. exp beta adrenergic receptor blocking agent/ OR exp Adrenergic beta-Antagonists/
11. (beta blocker$ OR BB OR acebutolol OR atenolol OR betaxolol OR bisoprolol OR carvedilol OR labetalol OR metoprolol OR nadolol OR nebivolol OR penbutolol OR pindolol OR propranolol OR sotalol OR timolol).mp.
12. exp aldosterone antagonist/
13. (aldosterone antagonist$ OR mineralocorticoid-receptor antagonist OR MRA OR eplerenone OR spironolactone).mp.
14. exp angiotensin receptor antagonist/
15. (angiotensin receptor blocker$ OR angiotensin receptor antagonist$ OR ARB OR azilsartan OR candesartan OR eprosartan OR irbesartan OR losartan OR olmesartan OR telmisartan OR valsartan).mp.
16. sodium-glucose co-transporter 2 OR SGLT2 OR SGLT2 inhibitor* OR sodium glucose adj6 inhibitor* OR (SGLT2 inhibitor*) OR sodium-glucose adj6 inhibitor* OR Sodium-Glucose Transporter 2 OR sodium glucose-cotransporter 2 OR sodium-glucose co-transporter$ OR sodium glucose-cotransporter$
17. (dapagliflozin OR empagliflozin).mp.
18 exp ivabradine plus metoprolol/ or exp ivabradine/ or exp carvedilol plus ivabradine/
19 (Omecamtiv mecarbil OR CK-1827452 OR Omecamtiv OR mecarbil)
20 (Vericiguat OR guanylate cyclase stimulator OR guanylate cyclase stimulator)
21. (Hydralazine-Isosorbide Dinitrate OR Hydralazine-Isosorbide adj6 Dinitrate OR Hydralazine Isosorbide Dinitrate OR Hydralazine adj6 Isosorbide adj6 Dinitrate)
22. OR/6-21
23. "randomized controlled trial".pt.
24. (random$ or placebo$ or single blind$ or double blind$ or triple blind$).ti,ab.
25. (retraction of publication or retracted publication).pt.
26. OR/23-25
27. "Quality of Life"/
28. (kansas city cardiomyopathy questionnaire or kccq)
29. ("minnesota living with heart failure questionnaire" or mlhfq)
30. ("quality of life" or qol or hrqol)

31. OR/27-30
32. 22 AND 31
33. (animals not humans).sh.
34. ((comment or editorial or meta-analysis or practice-guideline or review or letter or journal correspondence) not "randomized controlled trial").pt.
35. (random sampl$ or random digit$ or random effect$ or random survey or random regression).ti,ab. not "randomized controlled trial".pt.
36. 33 OR 34 OR 35
37. 26 NOT 36
38. (random$ or placebo$ or single blind$ or double blind$ or triple blind$).ti,ab.
39. RETRACTED ARTICLE/
40. OR/38-39
41. (animal$ not human$).sh,hw.
42. (book or conference paper or editorial or letter or review).pt. not exp randomized controlled trial/
43. (random sampl$ or random digit$ or random effect$ or random survey or random regression).ti,ab. not exp randomized controlled trial/
44. OR/41-43
45. 40 NOT 44
46. 37 OR 45
47. 5 AND 32 AND 46
48. limit 47 to (human and yr="2024-Current" and (adult <18 to 64 years> or aged <65+ years>))

***Cochrane Clinical Trial search***
**Date of search:** Feb 28 2023
#1 MeSH descriptor: [Heart Failure] explode all trees
#2 MeSH descriptor: [Cardiomyopathy, Dilated] explode all trees
#3 (heart failure or cardiac failure or cardiac insufficiency or cardiomyopath$):ti,ab,kw (Word variations have been searched)
#4 #1 or #2 or #3
#5 (LCZ696 or LCZ 696 or LCZ-696 or valsartan adj6 sacubitril or valsartan adj6 sacubitril or valsartan sacubitril or valsartan-sacubitril or sacubitril adj6valsartan or sacubitril valsartan or sacubitril-valsartan):ti,ab,kw
#6 MeSH descriptor: [Angiotensin-Converting Enzyme Inhibitors] explode all trees
#7 (angiotensin converting enzyme inhibitor or ACEI or ACEI or antagonist$ or inhibitor$ benazepril or captopril or enalapril or fosinopril or imidapril or lisinopril or moexipril or perindopril or quinapril or ramipril or trandolapril or zofenopril or alacepril or cilazapril or spirapril or delapril):ti,ab,kw (Word variations have been searched)
#8 MeSH descriptor: [Adrenergic beta-Antagonists] explode all trees
#9 (beta blocker$ or BB or acebutolol or atenolol or betaxolol or bisoprolol or carvedilol or labetalol or metoprolol or nadolol or nebivolol or penbutolol or pindolol or propranolol or sotalol or timolol):ti,ab,kw (Word variations have been searched)
#10 MeSH descriptor: [Mineralocorticoid Receptor Antagonists] explode all trees

#11 (aldosterone antagonist$ or mineralocorticoid-receptor antagonist or MRA or eplerenone or spironolactone):ti,ab,kw (Word variations have been searched)
#12 MeSH descriptor: [Angiotensin Receptor Antagonists] explode all trees
#13 (angiotensin receptor blocker$ or angiotensin receptor antagonist$ or ARB or azilsartan or candesartan or eprosartan or irbesartan or losartan or olmesartan or telmisartan or valsartan):ti,ab,kw (Word variations have been searched)
#14 MeSH descriptor: [Sodium-Glucose Transporter 2 Inhibitors] explode all trees
#15 (sodium-glucose co-transporter 2 or SGLT2 or SGLT2 inhibitor* or sodium glucose adj6 inhibitor* OR (SGLT2 inhibitor*) or sodium-glucose adj6 inhibitor* or Sodium-Glucose Transporter 2 or sodium glucose-cotransporter 2 or sodium-glucose co-transporter$ or sodium glucose-cotransporter$):ti,ab,kw (Word variations have been searched)
#16 (dapagliflozin or empagliflozin):ti,ab,kw
#17 MeSH descriptor: [Ivabradine] explode all trees
#18 (ivabradine plus metoprolol or ivabradine or exp carvedilol plus ivabradine):ti,ab,kw
#19 (Omecamtiv mecarbil or CK-1827452 or Omecamtiv or mecarbil):ti,ab,kw
#20 (Vericiguat OR guanylate cyclase stimulator OR guanylate cyclase stimulator):ti,ab,kw
#21 (Hydralazine-Isosorbide Dinitrate OR Hydralazine-Isosorbide adj6 Dinitrate OR Hydralazine Isosorbide Dinitrate OR Hydralazine adj6 Isosorbide adj6 Dinitrate):ti,ab,kw
#22 MeSH descriptor: [Quality of Life] explode all trees
#23 (kansas city cardiomyopathy questionnaire or kccq):ti,ab,kw
#24 ("minnesota living with heart failure questionnaire" or mlhfq):ti,ab,kw
#25 ("quality of life" or qol or hrqol):ti,ab,kw
#26 #22 or #25 or # 23 or #24
#27 #5 or #6 or #7 or #8 or #9 or #10 or #11 or #12 or #13 or #14 or #15 or #16 or #17 or #18 or #19 or #20 or #21
#28 #4 and #26 and #27
#29 human not animal
#30 #28 and #29
#31 #30 in trials
#32 #31 (limit from 2020)

***Cochrane Clinical Trial search***

**Date of search:** Feb 20 2024
#1 MeSH descriptor: [Heart Failure] explode all trees
#2 MeSH descriptor: [Cardiomyopathy, Dilated] explode all trees
#3 (heart failure or cardiac failure or cardiac insufficiency or cardiomyopath$):ti,ab,kw (Word variations have been searched)
#4 #1 or #2 or #3
#5 (LCZ696 or LCZ 696 or LCZ-696 or valsartan adj6 sacubitril or valsartan adj6 sacubitril or valsartan sacubitril or valsartan-sacubitril or sacubitril adj6valsartan or sacubitril valsartan or sacubitril-valsartan):ti,ab,kw
#6 MeSH descriptor: [Angiotensin-Converting Enzyme Inhibitors] explode all trees
#7 (angiotensin converting enzyme inhibitor or ACEI or ACEI or antagonist$ or inhibitor$ benazepril or captopril or enalapril or fosinopril or imidapril or lisinopril or moexipril or perindopril or quinapril or

ramipril or trandolapril or zofenopril or alacepril or cilazapril or spirapril or delapril):ti,ab,kw (Word variations have been searched)
#8 MeSH descriptor: [Adrenergic beta-Antagonists] explode all trees
#9 (beta blocker$ or BB or acebutolol or atenolol or betaxolol or bisoprolol or carvedilol or labetalol or metoprolol or nadolol or nebivolol or penbutolol or pindolol or propranolol or sotalol or timolol):ti,ab,kw (Word variations have been searched)
#10 MeSH descriptor: [Mineralocorticoid Receptor Antagonists] explode all trees
#11 (aldosterone antagonist$ or mineralocorticoid-receptor antagonist or MRA or eplerenone or spironolactone):ti,ab,kw (Word variations have been searched)
#12 MeSH descriptor: [Angiotensin Receptor Antagonists] explode all trees
#13 (angiotensin receptor blocker$ or angiotensin receptor antagonist$ or ARB or azilsartan or candesartan or eprosartan or irbesartan or losartan or olmesartan or telmisartan or valsartan):ti,ab,kw (Word variations have been searched)
#14 MeSH descriptor: [Sodium-Glucose Transporter 2 Inhibitors] explode all trees
#15 (sodium-glucose co-transporter 2 or SGLT2 or SGLT2 inhibitor* or sodium glucose adj6 inhibitor* OR (SGLT2 inhibitor*) or sodium-glucose adj6 inhibitor* or Sodium-Glucose Transporter 2 or sodium glucose-cotransporter 2 or sodium-glucose co-transporter$ or sodium glucose-cotransporter$):ti,ab,kw (Word variations have been searched)
#16 (dapagliflozin or empagliflozin):ti,ab,kw
#17 MeSH descriptor: [Ivabradine] explode all trees
#18 (ivabradine plus metoprolol or ivabradine or exp carvedilol plus ivabradine):ti,ab,kw
#19 (Omecamtiv mecarbil or CK-1827452 or Omecamtiv or mecarbil):ti,ab,kw
#20 (Vericiguat OR guanylate cyclase stimulator OR guanylate cyclase stimulator):ti,ab,kw
#21 (Hydralazine-Isosorbide Dinitrate OR Hydralazine-Isosorbide adj6 Dinitrate OR Hydralazine Isosorbide Dinitrate OR Hydralazine adj6 Isosorbide adj6 Dinitrate):ti,ab,kw
#22 MeSH descriptor: [Quality of Life] explode all trees
#23 (kansas city cardiomyopathy questionnaire or kccq):ti,ab,kw
#24 ("minnesota living with heart failure questionnaire" or mlhfq):ti,ab,kw
#25 ("quality of life" or qol or hrqol):ti,ab,kw
#26 #22 or #25 or # 23 or #24
#27 #5 or #6 or #7 or #8 or #9 or #10 or #11 or #12 or #13 or #14 or #15 or #16 or #17 or #18 or #19 or #20 or #21
#28 #4 and #26 and #27
#29 human not animal
#30 #28 and #29
#31 #30 in trials
#32 #31 (limit from 2023)

***Cochrane Clinical Trial search***
**Date of search:** Aug 10 2024
#1 MeSH descriptor: [Heart Failure] explode all trees
#2 MeSH descriptor: [Cardiomyopathy, Dilated] explode all trees
#3 (heart failure or cardiac failure or cardiac insufficiency or cardiomyopath$):ti,ab,kw (Word variations have been searched)
#4 #1 or #2 or #3

#5 (LCZ696 or LCZ 696 or LCZ-696 or valsartan adj6 sacubitril or valsartan adj6 sacubitril or valsartan sacubitril or valsartan-sacubitril or sacubitril adj6valsartan or sacubitril valsartan or sacubitril-valsartan):ti,ab,kw
#6 MeSH descriptor: [Angiotensin-Converting Enzyme Inhibitors] explode all trees
#7 (angiotensin converting enzyme inhibitor or ACEI or ACEI or antagonist$ or inhibitor$ benazepril or captopril or enalapril or fosinopril or imidapril or lisinopril or moexipril or perindopril or quinapril or ramipril or trandolapril or zofenopril or alacepril or cilazapril or spirapril or delapril):ti,ab,kw (Word variations have been searched)
#8 MeSH descriptor: [Adrenergic beta-Antagonists] explode all trees
#9 (beta blocker$ or BB or acebutolol or atenolol or betaxolol or bisoprolol or carvedilol or labetalol or metoprolol or nadolol or nebivolol or penbutolol or pindolol or propranolol or sotalol or timolol):ti,ab,kw (Word variations have been searched)
#10 MeSH descriptor: [Mineralocorticoid Receptor Antagonists] explode all trees
#11 (aldosterone antagonist$ or mineralocorticoid-receptor antagonist or MRA or eplerenone or spironolactone):ti,ab,kw (Word variations have been searched)
#12 MeSH descriptor: [Angiotensin Receptor Antagonists] explode all trees
#13 (angiotensin receptor blocker$ or angiotensin receptor antagonist$ or ARB or azilsartan or candesartan or eprosartan or irbesartan or losartan or olmesartan or telmisartan or valsartan):ti,ab,kw (Word variations have been searched)
#14 MeSH descriptor: [Sodium-Glucose Transporter 2 Inhibitors] explode all trees
#15 (sodium-glucose co-transporter 2 or SGLT2 or SGLT2 inhibitor* or sodium glucose adj6 inhibitor* OR (SGLT2 inhibitor*) or sodium-glucose adj6 inhibitor* or Sodium-Glucose Transporter 2 or sodium glucose-cotransporter 2 or sodium-glucose co-transporter$ or sodium glucose-cotransporter$):ti,ab,kw (Word variations have been searched)
#16 (dapagliflozin or empagliflozin):ti,ab,kw
#17 MeSH descriptor: [Ivabradine] explode all trees
#18 (ivabradine plus metoprolol or ivabradine or exp carvedilol plus ivabradine):ti,ab,kw
#19 (Omecamtiv mecarbil or CK-1827452 or Omecamtiv or mecarbil):ti,ab,kw
#20 (Vericiguat OR guanylate cyclase stimulator OR guanylate cyclase stimulator):ti,ab,kw
#21 (Hydralazine-Isosorbide Dinitrate OR Hydralazine-Isosorbide adj6 Dinitrate OR Hydralazine Isosorbide Dinitrate OR Hydralazine adj6 Isosorbide adj6 Dinitrate):ti,ab,kw
#22 MeSH descriptor: [Quality of Life] explode all trees
#23 (kansas city cardiomyopathy questionnaire or kccq):ti,ab,kw
#24 ("minnesota living with heart failure questionnaire" or mlhfq):ti,ab,kw
#25 ("quality of life" or qol or hrqol):ti,ab,kw
#26 #22 or #25 or # 23 or #24
#27 #5 or #6 or #7 or #8 or #9 or #10 or #11 or #12 or #13 or #14 or #15 or #16 or #17 or #18 or #19 or #20 or #21
#28 #4 and #26 and #27
#29 human not animal
#30 #28 and #29
#31 #30 in trials
#32 #31 (limit from 2024)

### Protocol

**Aim**
Evaluate the efficacy of pharmacotherapy in improving health-related quality of life in patients with heart failure with reduced ejection fraction using a network meta-analysis approach.
**Population**
Patients with heart failure with reduced ejection fraction (LVEF<50%).
Participants aged ≥18 years to 70 years.
Participants given the study drug as outpatients or after stabilization following hospitalization for HF.
Entire studies exclusively enrolling a specific sub-population (e.g., patients post-myocardial infarction, or patients with diabetes) are excluded.
Studies treating patients exclusively in the acute phase of heart failure are excluded.
No limit on background therapy.
**Intervention**
Digoxin
ARB
MRA
ACEi
β-blockers
Isosorbide dinitrate and hydralazine
Sacubitril/valsartan
Ivabradine
SGLT2 inhibitors
Vericiguat
Omecamtiv-mecarbil
**Comparison**
Control: Placebo
Interclass comparisons: any of the medications listed above. Intraclass comparisons are excluded.
**Outcome**
Primary and Secondary

- Health status outcomes measured by the Kansas City Cardiomyopathy Questionnaire (KCCQ) and the Minnesota Living with Heart Failure Questionnaire (MLHFQ)

**Methods**

**Databases**
This study will include three databases: MEDLINE (via ncbi.nlm.nih.gov), EMBASE (via ovidsp.ovid.com), and Cochrane Central (via www.cochranelibrary.com).

**Database search**
Searches will be performed between January 1st 1986 and October 1st 2022 in all databases. Subject headings, MeSH terms, and /or keyword searches were based on the search terms used in a previous network meta-analysis (Tromp and Ouwerkerk et al. 2021)[21]. In this study previously used outcome parameters (e.g. mortality) were replaced with parameters related to quality of life including the scores assessed by the MLHFQ and KCCQ tools.

**Eligibility criteria**
*Inclusion criteria*
Adult populations (≥18 years)
Randomized clinical trials
HFrEF (left ventricular ejection fraction [LVEF] <50%)
Enrolled in the outpatient setting or after stabilization following hospitalization for HF
Drug classes currently recommended or proven effective in treating HFrEF, including ACEi, ARB, β-blockers, Isosorbide dinitrate and hydralazine, Sacubitril/valsartan, Ivabradine, SGLT2 inhibitors, Vericiguat, Omecamtiv-mecarbil, MRA's.

*Exclusion criteria*
Studies where the entire population had a concomitant diagnosis likely affecting outcome (e.g., LV dysfunction post myocardial infarction)
Studies treating patients in the acute phase of HF
Comparisons within the same drug class except when the background therapy is different
Non-English language studies

**Screening**
The search will be carried out by RM and NS using search terms described previously for studies published between January 1st 1986 and January 27th 2023. References are collated in Mendeley and duplicates will be removed. We will upload references to Rayyan (https://www.rayyan.ai/) to screen titles and abstracts. Titles and abstract will be screened by RM and NS, blinded to each other's choices. Any conflicts will be discussed and solved by consensus. Articles left after this initial screening will be read by RM and NS, separately. Reasons for exclusion of articles will be recorded. Similarly, any conflicts will be discussed and solved by consensus.

**Data extraction**
Two study authors (RM and NS) will extract the data in a parallel and independent fashion on a dedicated standardized spreadsheet in Microsoft Excel. The spreadsheets will be compared between the two reviewers to ensure validity of data extraction. The following information will be extracted:
-Trial information: authors, study acronym, year, journal of publication
-Characteristics of the trial: yes/no for double blind, multi center, multi country, duration, missing data, selective reporting, imbalanced dropout, similar groups at baseline, treatment allocation blinding, allocation concealment and randomization
-Per treatment arm: total number of participants, mean age with SD, mean LVEF with SD, number of men, number of patients with NYHA class I/II and III/IV, diabetes, ischemic heart failure, and diabetes mellitus, number of patients on background therapy and background therapy of patients, dosing of interventions
-Per treatment arm outcomes mean or median KCCQ clinical summary score (CSS) [±SD/IQR], total symptom score (TSS)[±SD/IQR], and overall summary score (OSS)[±SD/IQR], and physical limitation score (PLS)[±SD/IQR] MLHFQ score [±SD/IQR] and number of participants discontinuing the drug.

**Risk of bias assessment**
We will use the Cochrane Risk of Bias tool to assess risk of bias (RM). This will be done independently by two investigators, discrepancies will be resolved through discussion and mutual agreement. Risk of bias will be presented as a figure, summarizing the risk per component of bias. To further assess

confidence in the network meta-analysis, we implemented the Confidence in Network Meta-Analysis (CINeMA) framework to assess (i) within-study bias, (ii) reporting bias, (iii) indirectness, (iv) imprecision, (v) heterogeneity, and (vi) incoherence (RM). To assess within study bias, we will use the Cochrane tool classifying as 'low', 'medium' and 'high' across 6-domains including (missing data, selective reporting, imbalanced dropout, similar groups at baseline, treatment allocation blinding, allocation concealment and randomization). We will use the average of all 6 domains to assess overall within-study bias – if more than 3 domains are considered high risk, the study was considered high risk, otherwise it is considered low risk. Reporting bias will be formally tested using the Egger test, which assesses the symmetry of funnel plots. When this is non-significant, we consider the risk of reporting bias as 'low'. Indirectness will be assessed based on published guidelines. To assess heterogeneity, we will calculate $I^2$ values (RM). The used model will be determined by the degree of heterogeneity, with random effects favored in the presence of heterogeneity ($I^2$ >30%). Incoherence will be assessed using a global test based on a random-effects design-by-treatment interaction model.

**Statistical analyses**

A frequentist random and fixed effects network and pairwise meta-analysis will be performed using the netmeta package in R. The QoL scores from studies using the KCCQ or MLHFQ will be standardized based on relevant clinical improvement conversion ranges established by Nassif et al. (2017).[26] Within this framework in the KCCQ a 5-, 10-, and 20-point change in the KCCQ overall summary score corresponds to a small, moderate, and large clinical change in patients' health status and in the MLHFQ small, moderate and large clinical changes corresponded to a change in overall score of −6.67 points, −10.41 points, and −17.90 points respectively. Results will be presented as a mean difference with 95% CI or a hazard ratio with 95% CI. Studies with no events in either arm will be excluded. Studies with events in one arm, but no events in the other will have a continuity correction applied to the zero arm of 0.5. Heterogeneity will be assessed using the $I^2$ values, which represent the amount of inconsistency in our networks. Exploiting the fact that some treatments are combinations of common components, an additive component network meta-analysis model will be used to evaluate the influence of individual components. This model assumes that the effect of treatment combinations is the sum of the effects of its components. Calculating additive treatment effects from multiple randomized trials provides useful estimates of combination therapies. Treatments will be categorized by drug group combination using a patient threshold of 50%. if >50% of patients received concomitant pharmacotherapy of interest, the treatment is described as a combination therapy.

**Sensitivity analyses**

We will undertake to conduct sensitivity analyses which will evaluate the length of follow up time on the validity of the results. Additionally, we will evaluate the impact of the dosages of the trial drugs achieved by trial groups on the main effect measure. We will evaluate the validity of the conversion of the MLHFQ scores to scores on the scale of the KCCQ by evaluating the ranking of treatments when ranked in the MLHFQ scale and ranked in the scale of the converted scores. Additionally we will evaluate
